# Automated Cell-free Protein Synthesis for Distributed Biomanufacturing

**DOI:** 10.1101/2025.10.15.25338083

**Authors:** Severino Jefferson Ribeiro da Silva, Mohammad Simchi, Quinn Matthews, Pouriya Bayat, Justin R. J. Vigar, Serena Singh, Idorenyin A. Iwe, Bárbara N. R. Santos, David Duplat, Paula Benítez-Bolivar, Krištof Bozovičar, Ryan Fobel, Aidan Tinafar, Seray Cicek, Yuxiu Guo, Fahim Masum, Lauren A. Cranmer, Masoud Norouzi, Renata P. G. Mendes, Cielo Léon, Laís C. Machado, Wentao Wu, Soheil Talebi, Alexander Klenov, Ashyad Rayhan, Jurandy J. F. de Magalhães, Carlos F. Narváez, Moiz Charania, Marcelo H. S. Paiva, Gabriel da Luz Wallau, Tony Mazzulli, David Sinton, Adriana Bernal, Camila González, Lindomar Pena, Keith Pardee

## Abstract

Recombinant proteins are fundamental to modern medicine, enabling research, diagnostics, and drug discovery, yet their access is often restricted by centralized manufacturing and cold-chain distribution. Decentralized, on-site biomanufacturing can enable research, provide resilience, and advance personalized medicine. Here, we introduce MANGO (MANufacturing on the GO), a purpose-built, open-source device designed for automated, benchtop cell-free protein synthesis and purification. Computer-controlled synthesis and purification are mediated through biomedical-grade pumps, valves, and a microchannel manifold, enabling rapid protein production directly at the point-of-use. To demonstrate MANGO’s utility, we produced and validated a diverse range of proteins, including nanobodies against SARS-CoV-2 and TNF, cloning enzymes, and diagnostic enzymes, with performance equivalent to their commercial counterparts. We further deployed MANGO in Brazil and Colombia to support patient trials for SARS-CoV-2 and dengue virus diagnostics. These results establish MANGO as a low-cost, portable platform that can democratize access to bioreagents in both high- and low-resource settings and, ultimately, expand participation in the bioeconomy.

## MAIN

Biotechnology is transforming medicine and accelerating advances in fundamental life sciences research and drug discovery. Yet, access to the proteins that underpin these innovations remains constrained. Most proteins are not readily available off-the-shelf; instead, accessing them requires substantial expertise and infrastructure for cell-based protein expression and purification^1–4^. Similar barriers exist for accessing variants of common proteins, and the rise of computational protein design is driving further demand for rapid prototyping of *de novo-* designed proteins^5, 6^.

When proteins are commercially available, centralized biomanufacturing offers economies of scale and provides widespread benefits to research. However, this production model has gaps that lead to unmet needs. Since protein biomanufacturing requires capital-intensive facilities and specialized expertise, production is mainly concentrated in a few global hubs and distributed to users through cold chain shipping **(Fig. 1a)**^2, 7, 8^. While this centralized model is effective, the supply chain can be fragile and, in many parts of the world, adds logistical barriers, cost, and delays that can limit access and research capacity.

**Figure 1.**
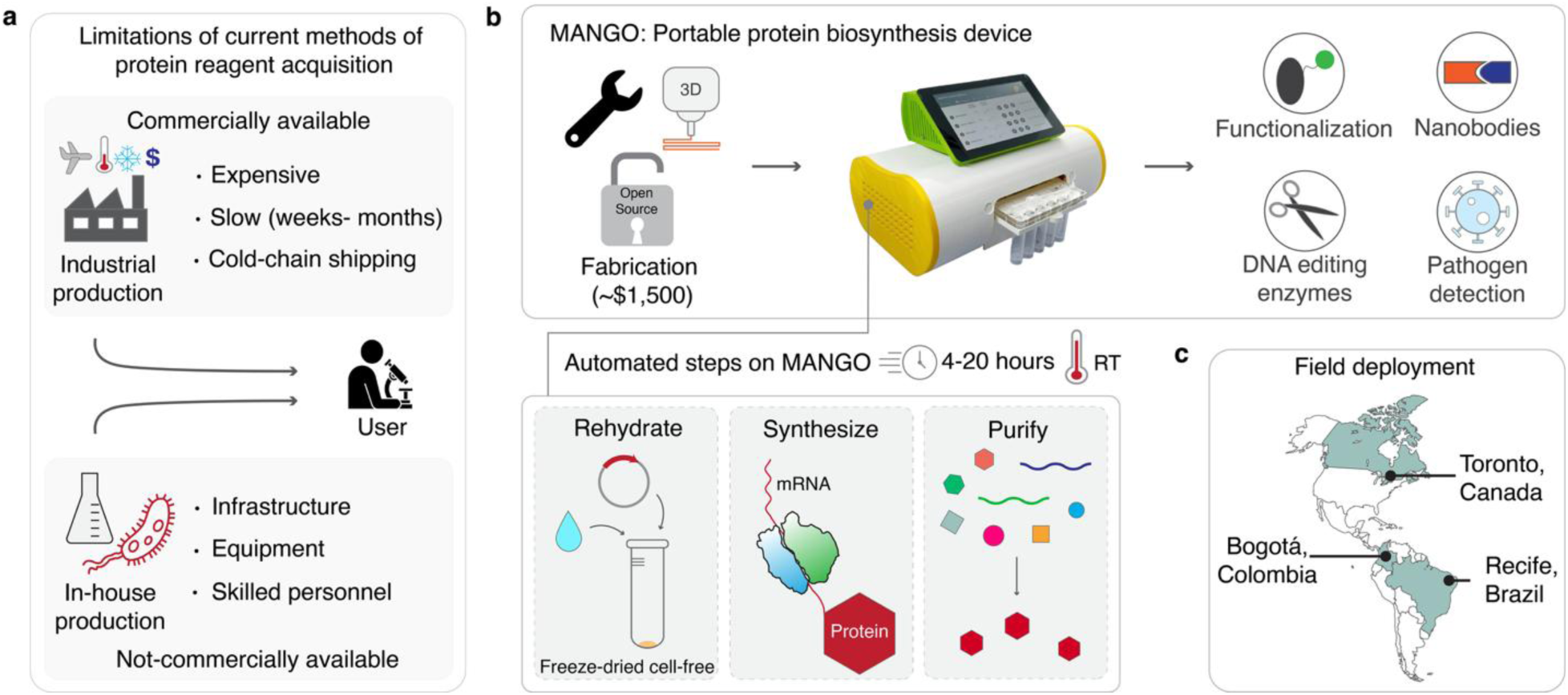
MANGO: A portable, low-burden biosynthesis device that brings on-demand protein production to the point-of-use. **(a)** Conventional protein production relies on centralized commercial facilities or well-equipped laboratories, requiring substantial infrastructure, skilled personnel, and extended timeframes (weeks to months). While this production system delivers significant advantages to those who can afford it, the associated costs, cold-chain transportation, and infrastructure demands create substantial barriers, restricting equitable access to bioreagents, especially in low- and middle-income countries (LMICs) and remote settings. This challenge is further compounded by limited commercial availability, where most proteins are not sold off-the-shelf, and only a small portion exists as catalog products. Similar challenges also arise for variants of common proteins, while the growth of computational protein design further increases the demand for rapid prototyping of *de novo*–designed proteins. **(b)** The MANGO (MANufacturing on the GO) platform enables distributed protein production using readily available components (estimated cost around $1,500 USD) and open-source designs. The system leverages freeze-dried, *E. coli* lysate-based CFPS reactions that are stable at ambient temperature and can be rehydrated and combined with template DNA at the point of use. With the MANGO in hand, the user primarily needs the manual instructions, the DNA-encoding template, and the FD-CFPS reactions on MANGO, which enables the easy production of valuable proteins anywhere with just the press of a button. MANGO automates protein synthesis and subsequent affinity chromatography purification, enabling the production of high-value products, including functionalized enzymes, nanobodies, DNA-modifying enzymes, and diagnostic enzymes. **(c)** Demonstrating true field readiness, MANGO was deployed to diagnostic laboratories in Recife, Brazil, and Bogotá, Colombia, where it produced RT-LAMP enzymes on-site for decentralized, molecular diagnostics. Once on-site in South America, the MANGO device produced enzymes within one day from FD-CFPS reactions, enabling the establishment of diagnostic programs for globally relevant pathogens.

These vulnerabilities are most evident during crises when reliance on centralized production may constrain access to essential products. Supply chain shocks during public health emergencies, such as the COVID-19 pandemic and other regional outbreaks^8–14^, have led to shortages of crucial bioreagents and limited laboratory capacity, hindering efforts for population-level surveillance and testing^11, 15, 16^. Similar bottlenecks also arise during natural disasters^17, 18^ and geopolitical conflicts^19, 20^, as well as in other scenarios that require urgent responses^9, 21^ (e.g., refugees and displaced persons).

Beyond acute crises, limited access to bioreagents is a daily challenge in low- and middle-income countries (LMICs), significantly reducing research capacity. In fact, the United Nations (UN) and the World Health Organization (WHO) have identified a lack of access to biotechnology as a critical challenge to development and capacity building^22, 23^. Our research groups in South America routinely face delivery times of up to four months for standard bioreagents, with costs two- to three-fold higher than in North America^24^. In LMICs, these constraints affect research that addresses local health priorities, including disease surveillance, fundamental life sciences research, and drug discovery, leaving regions in need of biotechnology solutions often unable to access or develop them.

Cell-free protein synthesis (CFPS) has emerged as a promising, low-burden platform for decentralized, on-site manufacturing that could improve access to bioreagents, diagnostic tools, and life-saving therapeutics^2, 25–34^. CFPS systems rely on cellular extracts containing transcription and translation enzymes, supplemented with energy sources and DNA templates that encode the protein of interest. This approach brings rapid protein production directly to the point-of-need^2, 27, 35^. Importantly, CFPS reagents can be lyophilized, allowing stable, ambient-temperature storage and simplified distribution, and can be prepared locally with basic microbiology equipment such as a flask shaker, centrifuge, and cell lysis tools^2, 36–39^.

CFPS has demonstrated the capacity to express a wide range of proteins, including those that are traditionally difficult to produce. These include proteins with disulfide bonds (e.g., granulocyte-macrophage colony-stimulating factor [GM-CSF])^34, 40, 41^, glycosylation^27, 28, 41, 42^, and proteins that are toxic to living cells^43^. Beyond the production of any single protein, on-demand and on-site biomanufacturing offers critical logistical advantages, echoing the benefits of on-site 3D printing of physical hardware for applications in daily life, health care, and military operations^44, 45^. By simply adding template DNA to a generic CFPS stockpile, users can produce a remarkably diverse set of protein products, enabling compact, yet broadly versatile biomanufacturing.

Despite these advantages, CFPS remains a laboratory-based method, and scaling its adoption and impact will require automation to de-skill use. Early advances in distributed manufacturing systems in medicine have already enabled the chemical synthesis of drugs using a refrigerator-sized instrument^46^, while the cell-based biosynthesis of small molecules has also been demonstrated using a portable bioreactor^47^. More recently, a suitcase-sized system has shown the potential for CFPS-based mobile protein production^48^.

This latter device, called BioMOD^48^, provided a compelling proof-of-concept of modular, portable CFPS biomanufacturing, showcasing the potential for field deployment. Built from commercially available components, such as syringe pumps, an incubated shaker, and a dialysis cartridge, housed in a Pelican™ case, BioMOD demonstrated both functionality and versatility. Yet, it was costly (estimated ∼ $6,000 USD), required skilled operators and manual intervention during production, and more closely resembled a breadboard prototype than a fully integrated device. These constraints limited its practical and scalable deployment.

To address the need for accessible, low-cost portable bioreagents production, we present MANGO (MANufacturing on the GO), a compact, fully integrated, user-friendly, and purpose-built benchtop device for automated CFPS-based manufacturing within hours at the push of a button **(Fig. 1b)**. Designed from the ground up for simple and low-cost assembly (∼$1,500 USD), MANGO is constructed from readily available materials and electronic components.

Fabricated from 3D-printed, laser-cut, and CNC-milled materials, and roughly the size of an American football, the open-source MANGO device allows researchers to synthesize and purify proteins on-demand in diverse settings. Proteins are synthesized on-device from fresh or freeze-dried *E. coli* lysate-based CFPS reactions and then purified via a fluidic manifold powered by pumps and valves. Users interact with MANGO through an onboard touchscreen with custom-designed software.

Following optimization of the fluidic system, we demonstrate MANGO’s versatility through a series of functional assays, benchmarking performance against commercial comparators at sites in Canada, Brazil, and Colombia to validate decentralized manufacturing. MANGO supports the production of a broad array of high-value proteins, including nanobodies targeting the tumor necrosis factor (TNF) and the SARS-CoV-2 spike protein, as well as the TNF cytokine itself, DNA-modifying enzymes for molecular biology (restriction enzymes, ligase), and diagnostic enzymes for detecting infectious diseases in regions of endemic infection (Brazil and Colombia). In reverse transcriptase loop-mediated isothermal amplification (RT-LAMP) assays, enzymes produced by MANGO achieved 100% concordance with the clinical gold standard RT-qPCR in detecting SARS-CoV-2 and dengue (DENV, serotypes 1 and 2) viruses from patient samples **(Fig. 1c**). These results establish MANGO as an accessible, portable platform for producing high-quality protein products and showcase its potential to enhance global access to biotechnology.

## RESULTS

### MANGO development and fabrication

With the goal of improving access to bioreagents, we have designed a low-cost and automated device for benchtop, cell-free protein synthesis and purification that can be thought of as a “Nespresso machine®” for protein production. The MANGO, which deskills the use of cell-free protein synthesis (CFPS), underwent three distinct generations of design, each addressing key engineering and technical challenges. The first generation was built as an open-layout prototype, utilizing peristaltic pumps to transfer fluids between reservoirs; however, this system was too large for practical deployment **(Fig. S1)**. The second-generation device was considerably smaller and utilized biomedical-grade dispense pumps connected by tubing inside a laser-cut acrylic housing **(Fig. S2).** While it worked well, the wiring and fluidic tubing-based design made it too complex for scalable assembly and introduced multiple potential points of failure.

Building on these lessons, the third and final MANGO design simplifies the system by integrating all electrical components into a custom, open-source driver board **(Fig. 2a, Fig. S3).** This board connects the biomedical-grade pumps and valves to a Raspberry PI microprocessor, streamlining automation and simplifying assembly **(Fig. S4)**. The system comprises a central control unit (Raspberry Pi and driver board) operated via an LCD touch screen interface that enables users to program and manage all steps of protein synthesis and purification through in-house, open-source software **(Fig. S5)**.

**Figure 2.**
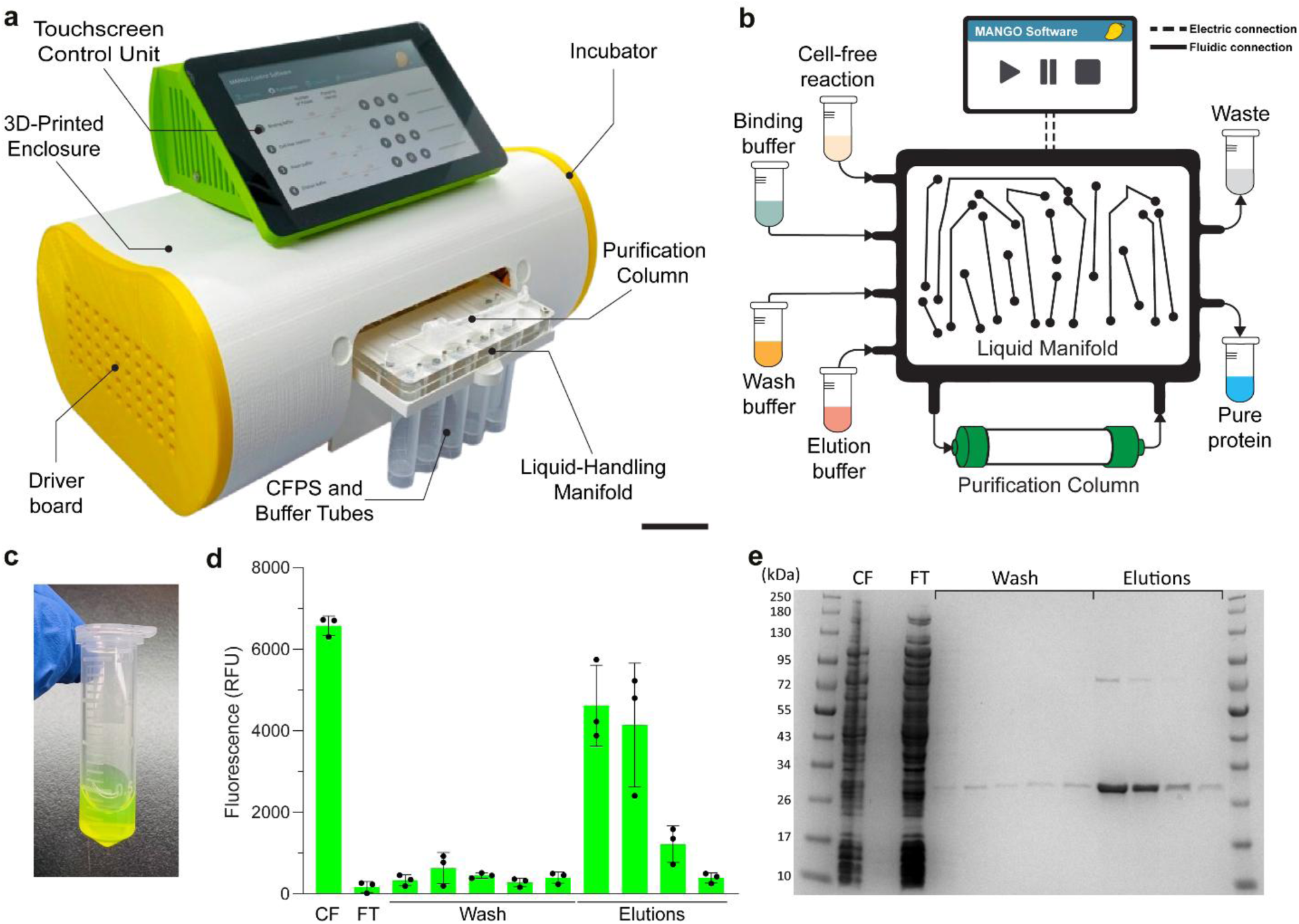
Automated protein synthesis and purification with MANGO. **(a)** The MANGO device includes a central control unit with an LCD touch screen, positioned on top of a 3D-printed curvilinear outer shell that houses and protects the incubator, electronics, and liquid-handling manifold. **(b)** The liquid-handling manifold displays the purification column and reagent pathways for cell-free protein synthesis (CFPS) and purification, from input to output tubes. Valves and pumps at the top control specific reagents or flow paths. **(c)** MANGO-produced and purified deGFP displayed a bright green color, serving as a visual indicator of successful expression and purification. **(d)** deGFP fluorescence was monitored throughout the purification process across three independent MANGO runs using a conventional plate reader. Data are presented as mean ± SD from three biological replicates (n = 3). **(e)** SDS-PAGE analysis of deGFP purification using MANGO, showing cell-free reaction (CF), wash, and elution fractions. The data represent one of three biological replicates, with similar results observed in the other two. **See Fig. S17 for details**. The molecular weight ladder (in kilodaltons) is displayed on the left. Abbreviations: FT, Flow through.

On the fluidics side, we adopted a similar approach to simplify assembly and enhance reliability by developing a reusable liquid handling manifold that consolidates fluid connections **(Fig. 2b)**. Biomedical-grade pumps and valves are mounted onto the clear acrylic manifold, creating a robust and compact liquid handling unit **(Fig. S6)**. The manifold also includes six screw-in ports for 5 mL tubes designated for the CFPS biosynthesis reaction, purification buffers, and waste products **(Figs. S6-7)**. Integrated within the fluid unit is a low-cost, in-house-manufactured purification column that can be packed with a purification matrix of choice.

All components are installed inside a 3D-printed curvilinear outer shell, which protects electrical and fluidic systems, enhancing MANGO’s portability and operational reliability (31.5×19.5×21 cm, ∼2.1 kg). The main shell houses the fluidic unit, and an incubator equipped with a temperature probe and fan, along with the driver board **(Fig. S8)**. This latter item, which serves as the central control hub for managing pumps and valves, enables precise and automated device operation. The manifold is conveniently accessed by sliding it out on steel rails through a magnetic-clamped door. Positioned atop the main shell is the control hub, providing a centralized and intuitive interface for system operation **(Fig. S9).** Power for the device is supplied through the back of the control unit, and with the correct adapter, the system can operate on worldwide power sources or even from a portable battery **(Fig. S10).**

### Device operation

MANGO streamlines protein production by integrating cell-free synthesis with automated expression and purification in a compact, self-contained system. Production begins by adding the CFPS reaction (fresh or freeze-dried, 0.5-2 mL final volume) along with template DNA encoding the protein of interest to the cell-free mixture tube (**Fig. S7**). The manifold system is then slid back into the MANGO, where it is incubated overnight. A built-in incubator maintains an optimal expression temperature for the product, which can range from room temperature to 37°C. To facilitate reaction aeration and CFPS mixing, a pulsatile pump delivers air into the reaction tube at a rate of 25 µL/sec (1 pumping pulse per second) **(Fig. S11)**^49^. The incubation duration and the aeration rate are fully adjustable via the graphical user interface (GUI).

While reaction duration can vary (>4 hours), a 16-hour synthesis time was typically selected to maximize yield. At the pre-programmed reaction end time, the system proceeds to automated purification using an on-board affinity purification column. The platform supports both immobilized metal affinity chromatography (IMAC) or Strep-Tactin-based purification. As with any protein expression process, aggregation can happen during CFPS. If protein aggregation occurs during synthesis, the supernatant can be clarified by manual filtration or by brief high-speed centrifugation before purification.

Prior to purification, the column is prepared by equilibration with binding buffer (40 column volumes). The CFPS reaction mixture is then passed through the column, where the affinity-tagged protein binds selectively to the resin. Weakly bound, off-target proteins are removed by sequential washing with binding buffer (16 column volumes) and wash buffer (80 column volumes), with waste routed to the waste tube (**Fig. S12**). Finally, the purified protein is released from the column with an elution buffer (16 column volumes) into a dedicated collection tube (**Fig. S12**). All fluidic operations, including reagent delivery, washing, and elution, are precisely managed by programmable pumps and valves under software control **(Figs. S5-7).** Key parameters, such as reaction and buffer volumes and flow rates, can be easily adjusted through the GUI, enabling customizable and reproducible purification protocols suitable for automated biomanufacturing workflows.

### MANGO characterization

To characterize the fluidic system, we first measured the dead volume (the amount of liquid retained within the channels, pumps, and valves of the manifold between the input and the outlet tubes) **(Fig. S12).** Dead volume is a critical parameter because it reflects the reproducibility of device fabrication, buffer usage, and the volumes needed for optimal protein yield and purity. Triplicate measurements of dead volume for binding, wash, and elution buffers, as well as the CFPS mixture, were performed across three different MANGO manifolds. The intra-device and inter-device replicate variation was found to be below 7%, reflecting consistent performance and reliable manufacturing **(Fig. S13)**.

Initial flow rate studies indicated that inconsistent packing density in purification columns significantly affected flow performance. To address this challenge, we standardized the column packing protocol using centrifugation at a constant speed **(Fig. S14, see Methods for details)**, yet we continued to observe variation in run-to-run flow rates.

To further assess the influence of individual components on flow performance, we assembled three MANGO devices with independent sets of pumps, valves, and purification columns **(Fig. S13).** We then measured the flow rate for each fluidic CFPS and buffer path using water **(Figs. S12 and S15)**, which revealed approximately 16% variation in flow rates between devices **(Fig. S15)**. To determine the influence of the purification column on flow rate variation, we then tested the system using the same column on each of the three devices, finding that columns contributed only 2-3% to system variability. Subsequently, we re-tested the manifolds using a single shared set of pumps, valves, and purification columns. Under these conditions, flow rate variation across devices dropped to below 7%, suggesting that pump manufacturing inconsistencies were the primary source of variation. This variation can be further minimized through standardized pump calibration during assembly.

### Validation with GFP as a model protein

Once the performance of the MANGO device was established, we used the expression of green fluorescent protein (GFP) as a model protein to prototype the system. Here, using a 1 mL CFPS reaction, His₆-tagged deGFP was expressed at ambient temperature for 16 hours with an aeration rate of one pulse per second (25 µL/sec). We assessed protein adsorption in the manifold by measuring the fluorescence signal of deGFP in the CFPS mixture before and after passing through the system without a purification column. A 10% reduction in fluorescence intensity was observed, indicating some protein adsorption within the manifold **(Fig. S16).** This effect could be minimized by using low protein-binding tubing.

We next expressed and purified deGFP in triplicate (1 mL CFPS reactions) using the MANGO device outfitted with a Ni–NTA affinity column **(see Methods for details).** As expected, fluorescence measurements revealed a strong GFP signal in the crude CFPS expression reaction and minimal background in the wash fractions, with most of the protein recovered in the first and second elution fractions **(Fig. 2c-e)**. Protein concentration measurements using the bicinchoninic acid assay (BCA) consistently reflected this pattern across all three runs, with protein yields exceeding 250 μg/mL and purity above 95% **(Fig. S17)**. Together, these data confirmed high column-binding efficiency, minimal loss during washing, and reproducible recovery of functional deGFP **(Figs. 2e, S17, and S18)**.

The MANGO device can be run in two modes. The first mode is semi-automated, allowing users to control the parameters of each step (synthesis, purification, etc.) in real-time through the GUI, which is used to develop protocols for each protein. Once a protocol is established, the MANGO can be operated in fully automated mode, eliminating the need for human intervention during the manufacturing process. To benchmark the automated purification capabilities of the MANGO platform, we expressed and purified deGFP using both manual and fully automated workflows. Electrophoresis, fluorescence, and protein quantification analyses showed strong agreement between manual and automated modes **(Fig. S19).**

### Protein co-expression and functionalization

Having established and validated GFP synthesis and purification on MANGO, we next tested the platform’s capability for *in situ* protein functionalization during CFPS. Here, deGFP was expressed with an N-terminal His₆ tag and a C-terminal AVI tag under standard MANGO reaction conditions. Simultaneous biotinylation was accomplished by simply adding BirA biotin-protein ligase and its cofactor mix to the CFPS reaction **(Fig. S20)**. As above, the synthesized protein was purified using Ni-NTA affinity chromatography on the MANGO device. To test the functionalized protein, the purified product was incubated with streptavidin-coated magnetic beads alongside a negative deGFP control that lacked an AVI tag. Beads were then washed and imaged, with the results confirming successful co-translational biotinylation of the AVI-tagged protein product using MANGO **(Fig. S20)**.

### On-demand production of functional nanobodies on MANGO

We next set out to demonstrate the device through a series of practical vignettes in three themes: i. therapeutic and research nanobodies, ii. an enzyme toolbox for cloning, and iii. high-value enzymes for molecular diagnostics. To begin, we targeted the automated production of nanobodies, which are small single-domain proteins with broad applications and advantages over IgG antibodies, including high stability and improved tissue penetration^50^.

To demonstrate MANGO as a tool for rapid on-site production of molecular binders, we expressed and validated two nanobodies (Nb), which are currently difficult to source through commercial channels **(Fig. 3a)**. The first, Nb21, targets the SARS-CoV-2 spike protein and has been validated for therapeutic use^51, 52^. We then produced a custom Nb targeting murine tumor necrosis factor (TNF) and the TNF cytokine itself, from design to *in vitro* validation, demonstrating MANGO’s ability to streamline drug discovery.

**Figure 3.**
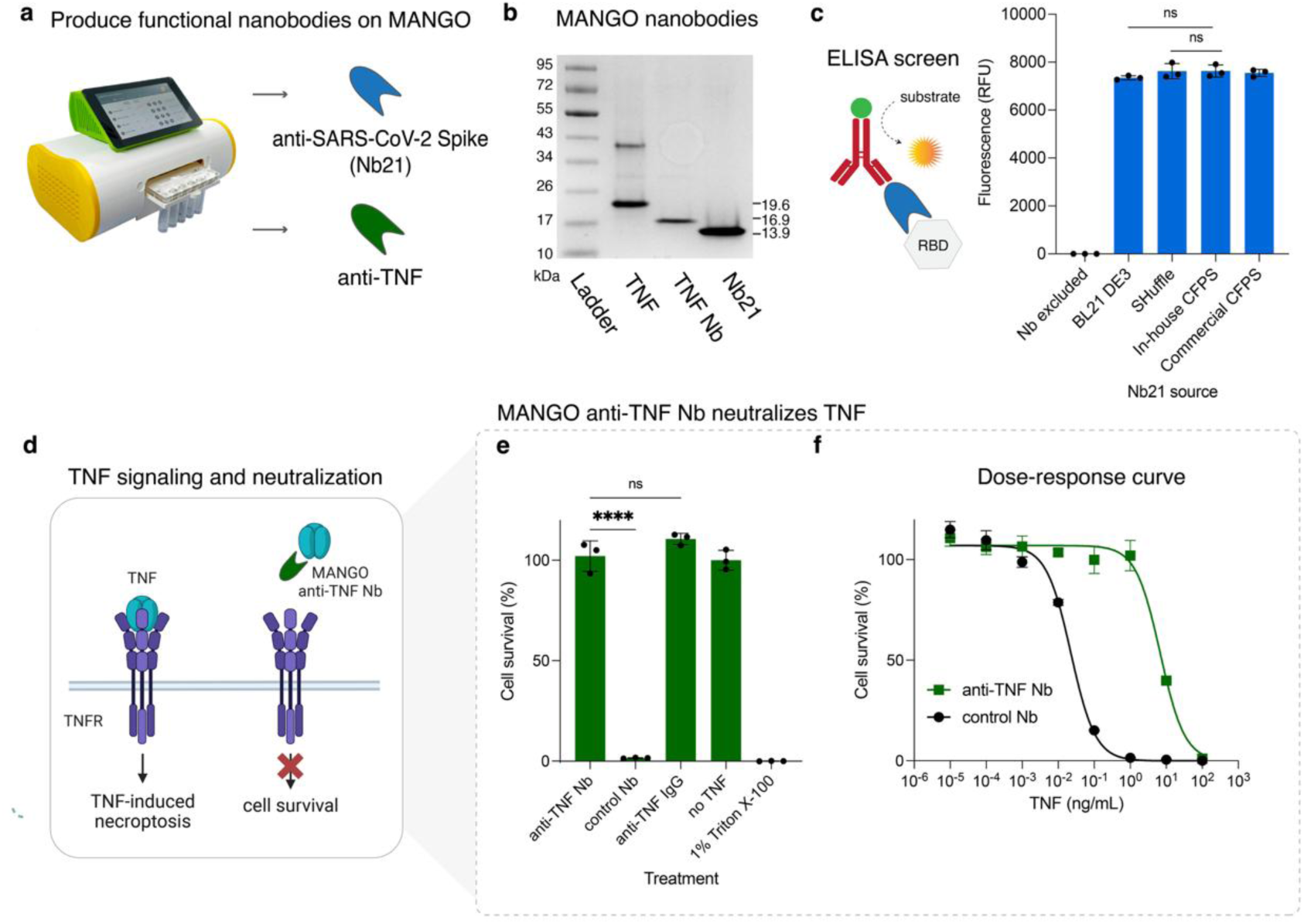
Production of functional nanobodies on MANGO. **(a)** Nanobodies targeting the SARS-CoV-2 spike protein and murine TNF, as well as the TNF itself, were produced on MANGO from CFPS reactions **(b)** After production, the products, including TNF (predominantly monomer with traces of dimer), TNF Nb, and anti-spike Nb (Nb21), were analyzed using 4-20% gradient SDS-PAGE and stained with ProBlue Safe, which confirmed their presence at the correct size. The molecular weight ladder (in kilodaltons) is shown on the left. **(c)** ELISA was performed against recombinant spike protein receptor-binding domain (RBD) using MANGO-produced anti-spike Nb21 as the primary antibody and HRP-conjugated anti-His₆ antibody as the secondary. Detection was performed using the QuantaBlu fluorogenic peroxidase substrate kit, and fluorescence intensity was measured in a conventional plate reader. **(d)** TNF neutralization assay was conducted using anti-TNF Nb (MANGO) and L929 cells, demonstrating **(e)** cell survival after preincubation of TNF (1 ng/mL) with both in-house and commercial antibodies. **(f)** Dose-response curves in the presence or absence of MANGO anti-TNF Nb. Luminescence was measured with a standard plate reader. Statistical significance was assessed using one-way ANOVA followed by Tukey’s multiple comparisons test. Data are shown as the mean of three technical replicates ± SD (ns p-value > 0.05, **** p-value < 0.0001). Abbreviations: TNF, tumor necrosis factor; TNFR, tumor necrosis factor receptor; Nb, nanobody.

Nb21, TNF Nb, and TNF were produced in CFPS reactions (1 mL, overnight at 22°C), after which the elution fractions were collected, quantified, and analyzed by gel electrophoresis, yielding products with endotoxin levels meeting standard guidelines (≤ 0.1 EU/mL) **(Fig. 3b, Fig. S21)**^53^. Nb21 was produced from two distinct DNA constructs off-device, one with an N-terminal His₆ tag and the other with a C-terminal His₆ tag, finding that the latter demonstrated better target binding and therefore proceeded with the C-terminally modified construct.

To confirm that nanobodies produced on MANGO retain functionality, a comparative binding screen was performed using Nb21 made on the device in parallel with traditional *E. coli-* based expression. Binding of Nb21 to recombinant spike protein was initially confirmed by dot blot analysis **(Fig. S22)**, followed by ELISA measurements using recombinant SARS-CoV-2 spike antigen and commercial HRP-conjugated anti-His₆ secondary antibody **(Fig. 3c, left)**. For cell-based expression, Nb21 was produced in two commonly used *E. coli* protein expression strains: BL21 (DE3) and SHuffle, a strain optimized for the expression of cysteine-rich proteins^54^. As expected, ELISA confirmed that MANGO Nb21 provided binding activity similar to that of nanobodies produced in BL21 DE3 and SHuffle strains **(Fig. 3c, right).** Overall, these findings confirm that nanobodies synthesized on the device are properly folded and capable of effective antigen interactions.

After establishing reliable nanobody production, we next integrated the MANGO platform into a drug development workflow. The ability of TNF Nb produced on MANGO to bind recombinant TNF and inhibit TNF-induced cytotoxicity was confirmed using the murine L929 fibroblast cell line **(Fig. 3d)**. L929 fibroblasts undergo necroptosis, a form of programmed cell death, when exposed to TNF in combination with actinomycin D, meanwhile, potent TNF inhibitors (such as nanobodies) mitigate the cytotoxic effects associated with TNF signaling^55, 56^. When cells were exposed overnight to 1 ng/mL TNF and a negative control nanobody, we observed near complete cell death, quantified by a luminescence-based ATP assay for cell viability **(Fig. 3e)**.

To test the capacity of MANGO TNF Nb to neutralize the cell-killing effects of TNF, we pre-incubated our in-house TNF Nb alongside a commercial anti-TNF IgG antibody, with results showing comparable cell survival **(Fig. 3e)**. Indeed, the cytotoxic dose 50% (CD_50_) values shifted from 0.02 to 7 ng/mL TNF upon treatment with in-house produced TNF Nb, reflecting a 350-fold decrease in toxicity **(Fig. 3f)**, and confirming that the MANGO-produced nanobody is biologically active. Additionally, we assessed the cytotoxic potential of recombinant TNF produced on MANGO and observed pronounced cell death, which was effectively rescued by antibody neutralization **(Fig. S23).** These proof-of-concept studies pave the way for the flexible, rapid, on-site production of nanobodies and potentially other antibody formats with broad applications.

### Scalable production of DNA assembly enzymes on MANGO

Enzyme-based tools for the assembly of DNA constructs are crucial for both fundamental and applied life sciences research, enabling the development of bespoke protein expression constructs, regulatory elements, sensors, and more^57, 58^. We sought to produce a toolkit of DNA assembly enzymes on MANGO to demonstrate how these bioreagents can be made more accessible **(Fig. 4a)**. Here, we selected four common cloning enzymes that typically require cold chain delivery: three restriction endonucleases (EcoRI, HindIII, and NcoI) for site-specific DNA cleavage and a ligase (T4 DNA ligase) for catalysis of phosphodiester bonds, enabling efficient DNA assembly.

**Figure 4.**
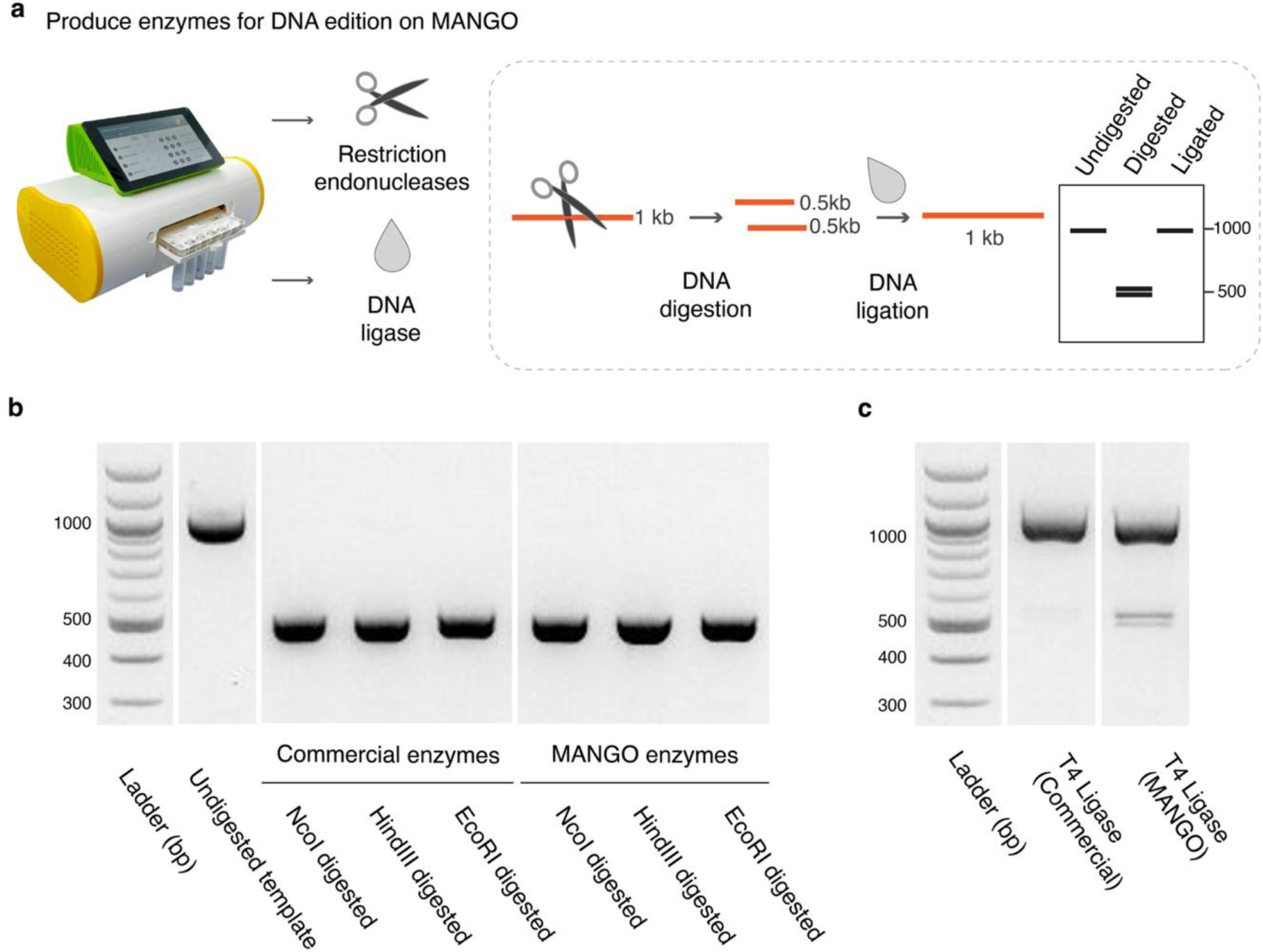
Scalable, on-demand production of core enzymes for DNA assembly on MANGO. **(a)** Schematic overview of the MANGO platform for on-demand synthesis of core DNA assembly enzymes, alongside the corresponding DNA digestion and ligation workflows. **(b)** Agarose gel electrophoresis comparing the DNA cleavage activity of commercial versus MANGO-produced restriction enzymes after 60 minutes of digestion. A 1 kb DNA substrate containing a central restriction site was used. Lane 1: DNA ladder; lane 2: undigested substrate; lanes 3–5: digestion by commercial enzymes; lanes 6–8: digestion by MANGO enzymes. Expected digestion products are approximately 500 bp fragments. **(c)** Agarose gel analysis of ligation activity comparing commercial and MANGO-produced T4 DNA ligase using a 1 kb DNA substrate pre-digested with EcoRI. Ligation products from commercial (lane 2) and MANGO (lane 3) enzymes are shown. The commercial ligase demonstrates higher efficiency, likely due to proprietary enzyme optimization; however, the MANGO ligase exhibits efficient and reliable ligation activity, making it suitable for DNA assembly workflows. Abbreviations: bp, base pairs. Kb, kilobase.

Using CFPS reactions (1 mL, overnight at ambient temperature), each enzyme fused to an N-terminal His₆ tag was produced and purified on MANGO, yielding soluble and functional products: HindIII, 149 µg; NcoI, 96 µg; EcoRI, 36 µg; and T4 DNA ligase, 107 µg. Here, all enzymes were successfully produced with high purity, confirming the efficiency and robustness of the MANGO platform **(Fig. S24)**.

With enzyme production protocols in place, we set out to characterize the activity of the enzymes produced on MANGO. To demonstrate the functionality of the enzymes, we designed a 1-kilobase (kb) DNA substrate containing restriction enzyme cut sites centrally positioned to evaluate cleavage efficiency **(Fig. S25, Table S1 for DNA sequence information)**. We began by assessing the activity of the restriction enzymes, defining their units based on the standard criteria: one unit is the amount of enzyme required to digest 1 µg of λ DNA in 1 hour at 37°C in a total reaction volume of 50 µL **(see Methods for details)**^59^. Using this definition, we determined the enzyme concentrations corresponding to one unit of activity: 1 nM (20 U/µL) for HindIII, 5 nM (20 U/µL) for EcoRI, and 50 nM (10 U/µL) for NcoI **(Fig. S25).**

Once optimal conditions for the restriction enzymes were established, we benchmarked the performance of the MANGO-produced restriction enzymes to their commercial counterparts. Both sets of enzymes were tested at a concentration of 100 nM, which, in all cases, provided 100% cleavage efficiency **(Fig. 4b, Fig. S25).** While it was not unexpected, the commercial enzymes operated with faster kinetics **(Fig. S25)**, which could possibly be attributed to their engineered, proprietary sequences and carefully optimized buffers.

Lastly, we assessed the activity of the MANGO-produced T4 DNA ligase. Ligation efficiency was tested at three incubation temperatures (16°C, 25°C, and 37°C) over 20-, 40-, and 60-minute intervals, with results showing comparable activity across all conditions **(Fig. S25)**. Building on these results, we next benchmarked the performance of the MANGO ligase against its commercial counterpart. Here, we simulated a cloning step with a 30-minute restriction enzyme digest followed by a 60-minute ligation. While the commercial ligase showed slightly higher performance, again possibly due to proprietary design, the enzyme set produced on-site by MANGO achieved efficient and reliable ligation **(Fig. 4c, Fig. S25)**. Taken together, these results confirm that MANGO-produced enzymes are effective molecular biology tools, providing a practical alternative to dependence on centralized biomanufacturing.

### Local production of diagnostic enzymes on MANGO

In our diagnostic efforts in South America^8, 13, 60–64^, we often experience logistical delays of 1-4 months for the arrival of key commercial enzymes^24^. It is through this lens that we sought to demonstrate MANGO for the on-site production of key bioreagents for detecting endemic and tropical mosquito-borne infections **(Fig. 5a,b)**. While RT-qPCR is the current gold standard for such molecular diagnostic assays^14, 65, 66^, the requirement for capital-intensive infrastructure (e.g., thermal cyclers) has limited its application in resource-constrained settings^67^. Isothermal amplification-based assays are increasingly filling this need in many settings. One such assay, loop-mediated isothermal amplification (LAMP), operates at a single temperature (60–65°C) and has emerged as a simple and powerful molecular diagnostic technique^68–71^. For pathogen DNA detection, LAMP requires only a strand-displacing polymerase, while RNA target amplification simply involves the addition of a reverse transcriptase^70^. In this work, these roles were fulfilled by expression of a large fragment of a strand-displacing polymerase from *Bacillus stearothermophilus* (Bst LF) and Moloney Murine Leukemia Virus reverse transcriptase (M-MLV)^70^, respectively.

**Figure 5.**
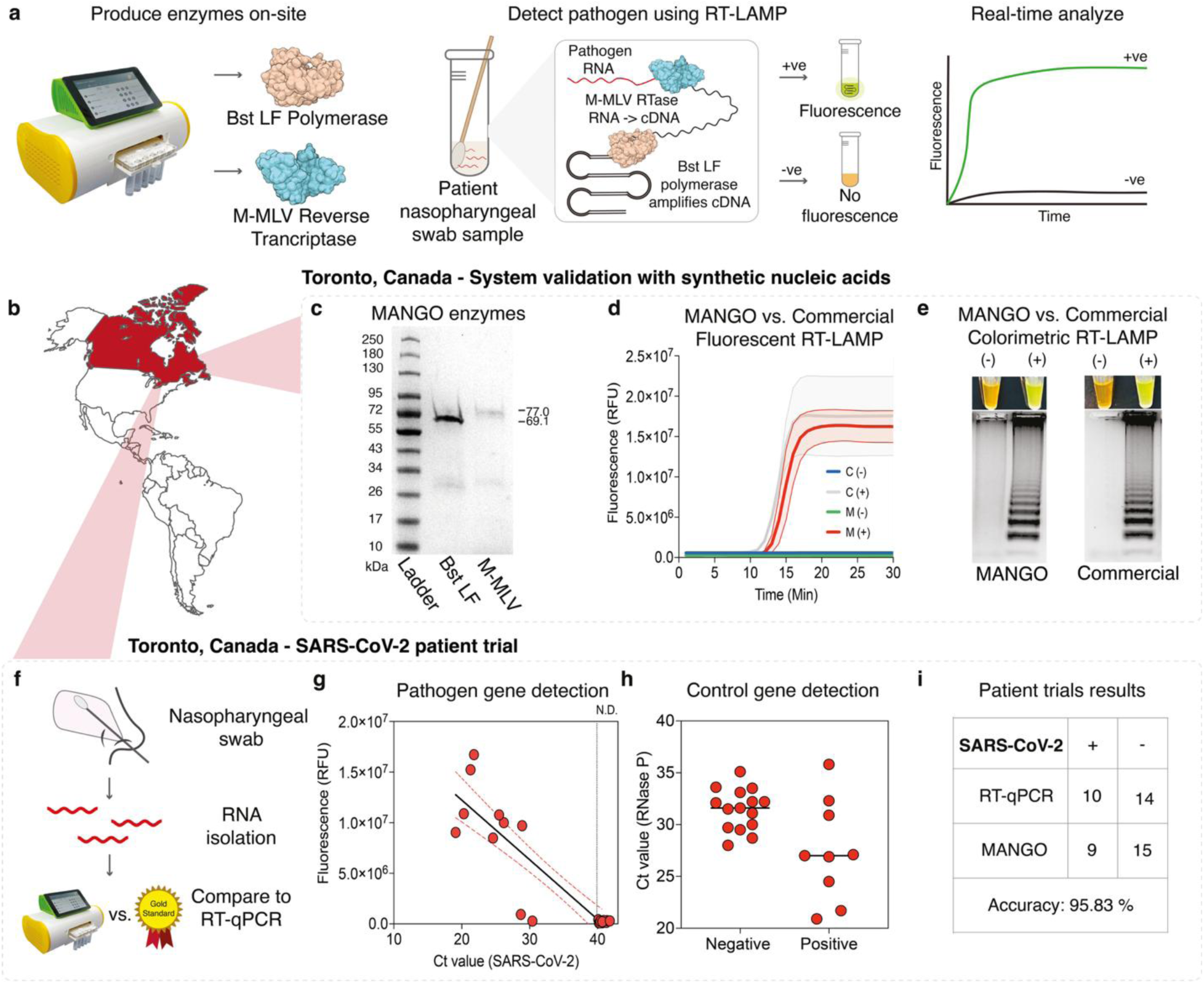
Infectious disease diagnostics with the MANGO device in Toronto, Canada. **(a)** This schematic explains the workflow for creating molecular diagnostic systems with enzymes produced through the MANGO platform. MANGO was used to produce and purify both Bst LF polymerase and M-MLV reverse transcriptase, which were subsequently incorporated into in-house RT-LAMP reactions for the detection of clinically relevant pathogens. Reactions were prepared using synthetic nucleic acids or nasopharyngeal swabs obtained from patients. Detection approaches included real-time fluorescence monitoring and visual endpoint assessment. In the fluorescence-based approach, amplicons were detected by adding 1X LAMP fluorescent dye, with measurements recorded at one-minute intervals using the RT-qPCR machine. For visual detection, SYBR Gold Nucleic Acid Stain (diluted 1:10) was applied to the tube caps before incubation. Following amplification, the tubes were mixed and examined under natural light. Positive reactions were indicated by a color change from orange to yellow, whereas negative reactions remained orange. **(b,c)** BstLF and M-MLV purified with the MANGO device were analyzed by SDS-PAGE (10–20% acrylamide) in experiments conducted in Toronto, Canada. The molecular weight ladder (in kilodaltons) is shown on the left. **(d)** Comparison of RT-LAMP performance between a commercial system and the in-house RT-LAMP system containing MANGO-produced enzymes using synthetic SARS-CoV-2 (nucleocapsid gene) RNA. Real-time fluorescence amplification was performed on a conventional qPCR instrument, with data presented as the mean ± SD (n = 3). The data represent technical replicates from a single representative experiment (three independent experiments produced similar results). **(e)** Similarly, we conducted an additional benchmark experiment to compare the performance of a commercial RT-LAMP kit with our in-house system, which contains MANGO-derived enzymes, using synthetic SARS-CoV-2 RNA and visualizing the results via colorimetric detection. Reactions were incubated in a conventional thermal cycler, and detection was based on a visible color change: a yellow color indicated a positive reaction, whereas an orange color indicated a negative reaction. The representative images shown were collected from one of three independent experiments. Together, these results demonstrate that the MANGO can produce enzymes with performance equivalent to their commercial counterparts. **(f)** Having established the diagnostic assay using on-site–produced reagents, we conducted a patient trial with 24 nasopharyngeal samples collected from suspected COVID-19 cases in Toronto, Canada. **(g)** Side-by-side testing of these samples was performed using RT-qPCR as the gold-standard comparator. Patient-derived RNA samples were tested using RT-LAMP, with an increase in fluorescence indicating successful amplification. Fluorescence measured at 30 minutes (y-axis) was plotted against the corresponding Ct values obtained from CDC RT-qPCR gold-standard assays (x-axis). **(h)** All RNA samples tested positive for RNase P (Ct values, y-axis) and in-house RT-LAMP results (negative or positive, x-axis; **see Fig. S29**), confirming their high quality. **(i)** Table summarizing the SARS-CoV-2 patient trial results and concordance with RT-qPCR, demonstrating an overall accuracy of 95.83%. **See Table S1 for detailed analysis**. Abbreviations: NTC, non-template control; Ct, cycle threshold; Min, minutes; RNase P, ribonuclease P; M, MANGO enzymes; C, commercial RT-LAMP kit.

Using CFPS reactions (1 mL, at ambient temperature), both Bst LF (His₈-tagged at the N-terminus) and M-MLV (His₆-tagged at the C-terminus) were successfully expressed and purified on MANGO using affinity-based chromatography. The products were then analyzed by electrophoresis and a colorimetric-based protein quantification assay, yielding 220 μg of Bst LF and 150 μg of M-MLV **(Fig. 5c, Fig. S26)**. Here, a 1 mL CFPS reaction on MANGO (CFPS input cost: ∼ $ 6.60 USD per product) yielded Bst LF and M-MLV in amounts sufficient for over 3,000 and 6,900 reactions, respectively. If purchased commercially, these quantities of enzyme would cost over $7,800 USD **(Fig. 5c, Appendix 1 for a cost breakdown).** After production, MANGO-derived enzymes were used to build an RT-LAMP kit on-site, and titration experiments were performed to optimize enzyme loading and reaction conditions before their use in downstream diagnostic applications.

With the enzymes and the diagnostic protocols in place, we then set out to benchmark our MANGO RT-LAMP kit against a commercial counterpart. To achieve this, we conducted a side-by-side comparison test to detect synthetic SARS-CoV-2 RNA using both fluorescent and colorimetric readouts **(see Methods for details)**. Reactions were incubated at 65 °C for 30 minutes, with fluorescence tracked in either real-time or visualized at the endpoint using SYBR Green–based colorimetric detection. Tracking amplicon generation fluorescently, we observe that in-house and commercial assays exhibit similar detection times (11–12 min), comparable rates of signal increase, and reach saturation with similar fluorescence intensity **(Fig. 5d).** Consistent with these results, both colorimetric detection and electrophoresis analysis also reliably showed comparable performance between the in-house and commercial systems **(Fig. 5e)**, demonstrating that enzymes produced with the MANGO device provide equivalent performance to their commercial counterparts.

Having confirmed the activity of Bst LF and M-MLV produced on MANGO, we next sought to demonstrate how MANGO enzymes can be applied in establishing disease diagnostic programs and on-site patient testing. Here, using MANGO-produced enzymes, we performed RT-LAMP on RNA extracted from 24 nasopharyngeal samples collected during the COVID-19 pandemic in Toronto, Canada, to detect SARS-CoV-2 RNA in parallel with the U.S. CDC gold-standard RT-qPCR assay^72^ **(Fig. 5f)**.

In a double-blinded study format, in-house RT-LAMP reactions were incubated at 65 °C, and fluorescence signal intensity was monitored every minute during a 30-minute incubation. Assays were conducted on patient samples with Ct values for SARS-CoV-2 (nucleocapsid gene) ranging from 19.1 to 30.4, along with RNase P (Ct values 20.9–35.8) as an endogenous human control to verify RNA quality and integrity **(Fig. 5g,h)**. Compared to RT-qPCR, we found a 95.83% (CI, 78.88% to 99.89%) diagnostic accuracy for RT-LAMP using MANGO enzymes **(Fig. 5i, Table S1)**. Notably, our in-house RT-LAMP assay showed 100% diagnostic accuracy in samples with Ct values ≤ 30, while a single false-negative outcome was observed in a sample with a Ct value > 30. This result was expected, while LAMP is an excellent option for diagnostics in low-resource settings, RT-qPCR is known to provide often more sensitive detection in direct comparisons for SARS-CoV-2^73^. With that in mind, these results demonstrate that MANGO can produce high-fidelity enzymes that provide results comparable to RT-qPCR, while also offering the advantage of being a low-burden, field-deployable option for decentralized, on-site diagnostics.

### MANGO deployment in South America enabled decentralized enzyme production for endemic disease diagnostics

A key goal in developing MANGO was to create a portable device suitable for global deployment, and so, through this lens, we next tested the MANGO at two sites in South America (Brazil and Colombia). Here, we repeated our validation of the RT-LAMP diagnostic enzymes (Bst LF polymerase and M-MLV reverse transcriptase) with a focus on the local need to detect SARS-CoV-2 and dengue virus (DENV). As in Toronto, enzymes were produced on-site and validated with patient samples (n=46) using CDC gold-standard RT-qPCR comparator assays **(Fig. 6a,b)**. To demonstrate the potential of accessible hardware, we also used our previously published FluoroPLUM^13, 63^ to enable high-capacity diagnostic testing (384-well plate).

**Figure 6.**
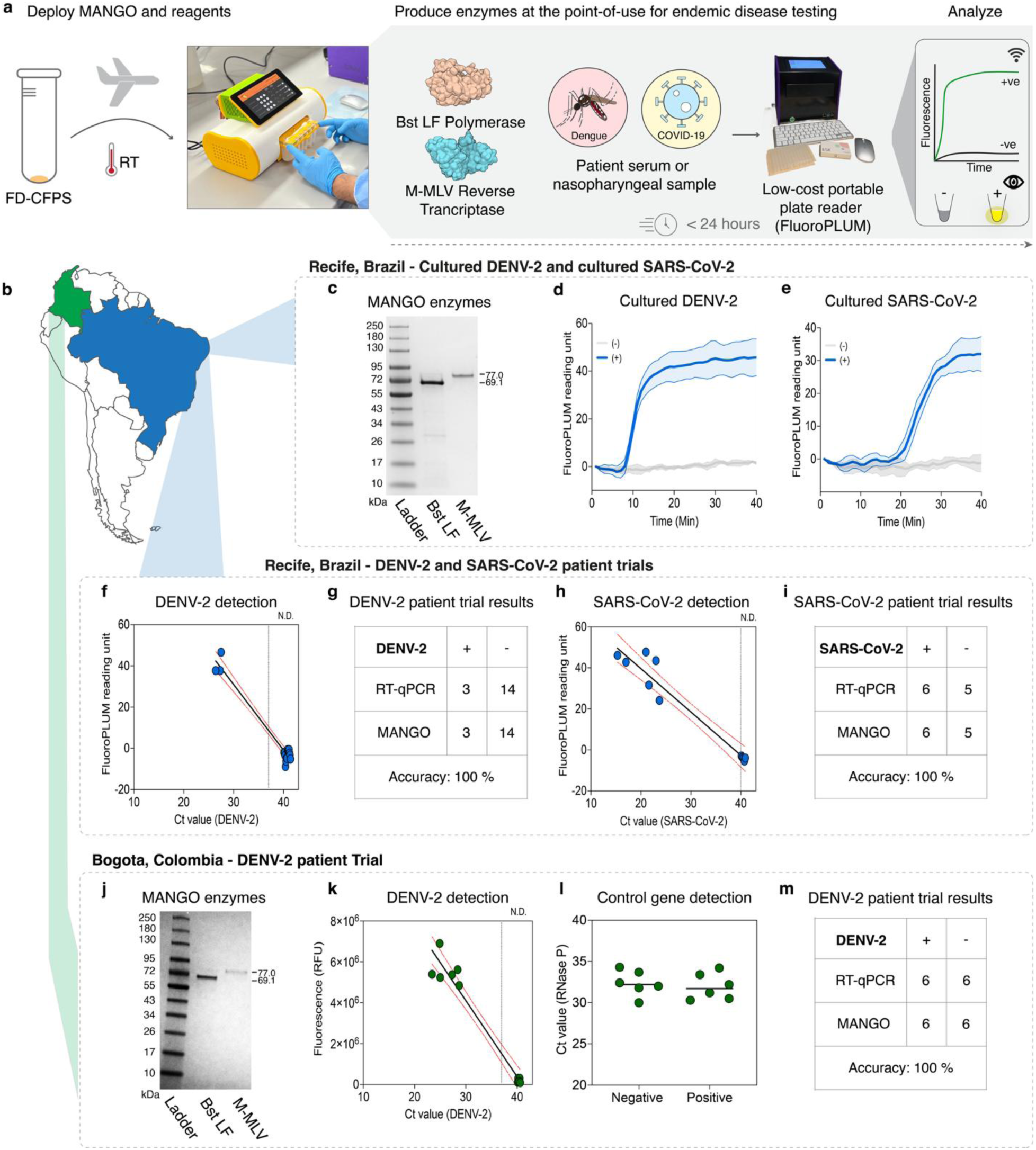
MANGO deployment in South America enabled on-site enzyme manufacturing and patient trials for dengue and SARS-CoV-2 viruses. **(a)** Schematic pipeline used to deploy MANGO and FluoroPLUM in South America, enabling on-site protein manufacturing and supporting the establishment of disease diagnostic programs in areas of endemic infection. Here, CFPS reactions were freeze-dried, vacuum-packed, and then transported from Toronto, Canada, to Brazil and Colombia at ambient temperature. **(b,c)** Once on site in Recife, Brazil, the initial field deployment location, FD-CFPS reactions were rehydrated. Bst LF and M-MLV enzymes were then synthesized overnight and purified using MANGO. After purification, both products were analyzed by SDS-PAGE (10–20% acrylamide), confirming their successful production. The molecular weight ladder (in kilodaltons) is displayed on the left. **(d,e)** With the enzymes and protocols established, the in-house RT-LAMP assays were initially tested using RNA isolated from cultured DENV-2 and SARS-CoV-2 viruses. SYTO-9 (at a final concentration of 10 μM) was used to visualize the results of the in-house RT-LAMP reactions. Real-time fluorescence detection was conducted on FluoroPLUM for 40 minutes with data presented as mean ± SD, n = 3. Data represent technical replicates from a single representative experiment (three independent experiments produced similar results). **(f)** Having confirmed the ability to detect targets under controlled settings, the in-house RT-LAMP functionality was tested against two panels of clinical samples in a national reference laboratory for viral infection diagnosis in Recife, Brazil. The first patient trial focused on the detection of DENV-2, and the results were compared to those obtained using the CDC RT-qPCR, which serves as the gold standard for comparison. Patient RNA samples were analyzed using RT-LAMP, where increasing fluorescence signals indicate successful amplification. The fluorescence measured (shown on the y-axis) was plotted against the corresponding Ct values obtained from CDC RT-qPCR (x-axis). **(g)** In-house diagnostic tests achieved a diagnostic accuracy of 100%. **See Table S2 for detailed analysis**. **(h)** Following these tests, an additional patient trial was performed to detect SARS-CoV-2 in nasopharyngeal samples. Patient-derived RNA samples were analyzed using RT-LAMP, with increasing fluorescence signals indicating successful detection and amplification. Fluorescence values recorded (y-axis) were then plotted against the corresponding Ct values determined by the CDC RT-qPCR assay (x-axis). **(i)** In-house diagnostic tests achieved a diagnostic accuracy of 100%. **See Table S3 for detailed analysis. (j)** Following the successful patient trial in Brazil, we deployed MANGO to Bogotá, Colombia, to again manufacture diagnostic enzymes on-site and support immediate patient testing. FD-CFPS reactions, prepared in Toronto, Canada, were shipped at ambient temperature and arrived ready for use. Within just 24 hours of setup, these reactions were rehydrated and used to produce both Bst LF and M-MLV directly on MANGO, demonstrating the platform’s ability to provide functional enzymes in resource-limited settings, as confirmed through SDS-PAGE (10–20% acrylamide) analysis. **(k-m)** With the inputs and protocols in place, a patient trial focused on the detection of DENV-2 was performed, and the results were compared to those obtained with the CDC RT-qPCR, serving as a gold standard. Patient RNA samples were analyzed with RT-LAMP, where increasing fluorescence signals indicate successful detection and amplification. The fluorescence measured (shown on the y-axis) was plotted against the corresponding Ct values obtained from CDC RT-qPCR (x-axis). In-house diagnostic tests achieved a diagnostic accuracy of 100%. **See Table S5 for detailed analysis.** In addition, all samples were positive for RNase P (RT-qPCR) and ACTB (RT-LAMP), confirming the integrity and high quality of the RNA samples. Abbreviations: NTC, non-template control; Ct, cycle threshold; Min, minutes; RNase P, ribonuclease P.

### Field deployment to Recife, Brazil

We first deployed MANGO in Recife, Pernambuco, Brazil—a hyper-endemic area for globally relevant mosquito-borne infections (Zika, dengue, and chikungunya), where the need for rapid, low-burden, decentralized molecular diagnostics is critical^74–76^. Within 24 hours of MANGO setup, freeze-dried (FD) CFPS reactions (1 mL, shipped from Toronto, Canada) enabled rapid enzyme production, yielding Bst LF (175.6 µg) and M-MLV (94 µg) after overnight incubation at ambient temperature with enzyme purity above 90% **(Fig. 6c, Fig. S27**).

With the enzymes and protocols in place, we began testing cultured DENV-2 and SARS-CoV-2 viruses to validate our in-house RT-LAMP reactions. Here, on-site in South America, reactions were monitored using the portable FluoroPLUM reader^13, 63^ at 65 °C for 30 minutes, with results confirming the detection of dengue serotype 2 (DENV-2) amplicons at ∼10 minutes and SARS-CoV-2 targets at ∼20 minutes **(Fig. 6d-e).**

Mosquito surveillance has become an essential addition to clinical diagnostics in public health efforts, playing a crucial role in monitoring viral circulation and ports of entry^77, 78^. To meet this need, we demonstrated our made-in-Brazil diagnostic assay by detecting DENV-2 in virus-spiked *Aedes aegypti* mosquito samples, the primary vector for dengue and other pathogens like Zika, chikungunya, and yellow fever viruses^79, 80^. DENV is a mosquito-borne pathogen that has been a major public health challenge in Brazil for over three decades^81^. Here, crude lysates of uninfected *Ae. aegypti* were spiked with cultured DENV-2 virus to achieve either a high viral load (1 × 10^6^ TCID_50_/mL) or a low viral load (1 × 10^3^ TCID_50_/mL)^82^, then directly used as templates in our in-house RT-LAMP assay. As expected, fluorescence signals showed successful detection of DENV-2 at both viral loads, consistent with the results obtained in parallel with the RT-qPCR **(Fig. S28)**.

After demonstrating pathogen detection in controlled conditions, we advanced to test the diagnostic system with patient samples. Working at a national reference laboratory for viral infections at Fiocruz in Recife, Brazil, we conducted two patient trials to evaluate the performance of our in-house RT-LAMP diagnostics in comparison to the CDC RT-qPCR assay^72, 83^. We first focused on detecting DENV-2. Here, in-house RT-LAMP reactions were incubated at 65 °C in the FluoroPLUM reader, with fluorescence signal intensity recorded every minute over a 30-minute incubation period. For the double-blind DENV-2 trial, RT-LAMP matched the positive (3 samples) and negative (14 samples) results found by RT-qPCR **(Fig. 6f, Fig. S29)**, resulting in a diagnostic accuracy of 100% (95% CI, 80.49% to 100%) **(Fig. 6g, see Table S2)**. Ct values for the target DENV-2 sequence (E gene)^83^ ranged from 26.4 to 27.5, along with RNase P, with Ct values ranging from 27.6 to 34.6 **(Fig. S29).**

With the high concordance observed between our in-house RT-LAMP test and RT-qPCR results, we advanced to a second patient trial to assess whether this diagnostic performance was maintained across a set of COVID-19 nasopharyngeal samples. For this purpose, we carried out in-house RT-LAMP assays on 11 RNA samples extracted from patient specimens collected in Recife, Brazil, during the COVID-19 pandemic. In parallel, all samples were tested side-by-side using the CDC RT-qPCR assay as a gold standard comparison^83^. Assays used samples with Ct values (nucleocapsid gene) ranging from 15.3 to 23.7, with RNase P Ct values ranging from 26.6 to 32.1 **(Fig. 6h, Fig. S29)**. Here, our in-house RT-LAMP assay accurately identified all positive (6) and negative (5) samples in a double-blinded format, achieving 100% (95% CI, 71.51% to 100%) diagnostic accuracy, consistent with RT-qPCR results **(Fig. 6i, Table S3)**.

### Field deployment to Bogotá, Colombia

Building on the success of the patient trials onsite in Brazil, we then deployed MANGO to Bogotá, Colombia, to again produce the RT-LAMP enzymes for detecting both dengue serotype 1 (DENV-1) and DENV-2, which are endemic to the region and have been associated with recent large-scale outbreaks^84, 85^. As before, FD-CFPS reactions (prepared in Toronto, Canada) were transported at ambient temperature to Bogotá, Colombia. Here, FD-CFPS reactions (1 mL) were rehydrated and run on MANGO, yielding 191 μg Bst LF and 81.5 μg M-MLV and purity above 90% **(Fig. 6j, Fig. S30)**.

With these locally produced diagnostic reagents in place, we next validated our in-house RT-LAMP using synthetic, *in vitro* transcribed RNA (DENV-1 and DENV-2) targets in preparation for patient sample testing **(Fig. S31)**. RT-LAMP reactions were monitored at 65 °C for 30 minutes, with results confirming the detection of both DENV (1 and 2) targets at approximately 10 minutes **(Fig. S31)**.

Having confirmed the activity of our enzymes, we began by testing our in-house RT-LAMP assay for DENV-1 using six clinical samples from patients suspected of having arbovirus infections. RT-LAMP reactions were performed at 65 °C, and fluorescence intensity was monitored at one-minute intervals over 30 minutes. Of these, three were confirmed positive for DENV-1 (NS5 gene) by CDC RT-qPCR^83^, with Ct values ranging from 18.4 to 25.7, and three were negative for DENV-1 **(Fig. S32)**. Here, the in-house RT-LAMP test demonstrated 100% (95% CI, 54.07% to 100%) diagnostic accuracy compared to the CDC RT–qPCR assay **(Fig. S32, Table S4)**. In addition to the DENV-1 genome target, all samples yielded positive results when tested for the beta-actin (ACTB) and RNase P housekeeping genes, confirming the RNA quality of the clinical samples **(Fig. S32).**

Encouraged by these results, we advanced to validate our in-house RT-LAMP assay targeting DENV-2. As in our work done in Brazil, a panel of serum samples (n=12) was tested side-by-side using our in-house RT-LAMP in comparison to the CDC RT-qPCR assay^83^. The tested samples exhibited Ct values ranging from 23.4 to 28.7, with RNase P values ranging from 30.0 to 34.3 **(Fig. 6k,l)**. Consistent with our previous findings, our RT-LAMP assay correctly identified all 6 positive and 6 negative samples under a double-blind format, achieving a diagnostic accuracy of 100% (95% CI, 73.54% to 100%), in agreement with the RT-qPCR results **(Fig. 6m, Table S5).** Together, these results highlight the practical utility of the MANGO platform in enabling on-site protein biomanufacturing and molecular diagnostics across diverse settings, ranging from national reference laboratories to low-resource settings.

## DISCUSSION

Emerging molecular tools in life sciences research and human health enable rational design and high precision, driving transformative advances in medicine. Yet the proteins underpinning this revolution (enzymes, antibodies) are costly, reliant on cold-chain logistics, and the capacity for their production is currently limited to settings with capital-intensive infrastructure and expertise. Whether it is access to fundamental bioreagents for life sciences research, enzymes for local diagnostics, or even prototyping for the coming revolution in artificial intelligence-driven protein design, accessible protein production is a deciding factor in local research capacity. While LMICs are especially impacted by these limitations, where high import fees, tariffs, and delays in customs clearance lead to 2-3 times higher costs and shipping delays of 1-4 months^24^, secure access to these tools is becoming increasingly important in all research settings as funding constraints grow.

Seeking to address this global challenge, we have developed and field-validated MANGO, a portable, low-burden, open-source device designed for benchtop, small-batch protein production. MANGO integrates all necessary steps for CFPS into a compact, user-friendly device, simplifying operation and enabling users to produce high-quality, functional protein products within hours by simply pressing a button.

To enable such portable biomanufacturing, we combined two core technologies: CFPS reactions that can be stored at ambient temperature for synthesis capacity that can be easily distributed, and MANGO to de-skill production. We first demonstrated MANGO’s utility for on-site protein production for life sciences research and biotechnology applications, where rapid access to diverse, functional proteins is crucial. Using MANGO, we successfully expressed nanobodies, enzymes for DNA manipulation, and high-value diagnostic enzymes at a fraction of the typical cost, providing a fast and flexible alternative to commercial sourcing. All MANGO-derived products exhibited performance and fidelity comparable to their commercially sourced counterparts **(Figs. 3-6**), supporting their suitability for research applications.

Recognizing that real-world challenges remain before global deployment, it is essential to ensure the consistent and reliable production of high-quality proteins with MANGO. In this study, we performed extensive quality assessments across sites and batches, demonstrating that MANGO-derived products exhibit reproducibility. To demonstrate MANGO’s global applicability and field readiness, we deployed units to infectious disease research and testing laboratories in Brazil and Colombia, settings where access to molecular reagents can be limited and timely diagnostics are critical^9, 24, 60^. Within a day of arrival, MANGO produced functional enzyme batches from rehydrated FD-CFPS reactions ($20 USD cost of goods for synthesis and purification), providing enzymes for over 3,000 RT-LAMP reactions at $0.27 per test, compared to over $1.40 per test for its commercial counterpart **(Appendix 1)**^8^. During on-site patient trials, MANGO-produced RT-LAMP enzymes showed high concordance with RT-qPCR gold-standard assays, achieving 100% accuracy **(Fig. 6, Tables S1-5)**. These demonstrations provided proof-of-concept for MANGO to address critical barriers to bioreagent access by enabling on-site protein production, reducing dependence on capital-intensive infrastructure, domain expertise, or complex logistics. In daily practice, this could provide researchers with access to on-demand bioreagents at a low cost, thereby lowering barriers imposed by limited budgets. We envision researchers having access to a companion device, like the MANGO, to provide on-demand, custom production. As evidenced by the rise in the accessibility and affordability of DNA synthesis, we anticipate that democratizing protein production will similarly accelerate what is possible and foster global access to the bioeconomy.

There are also potential strategic applications of the MANGO device. When paired with stockpiled FD-CFPS and DNA libraries, the MANGO could boost local capacity, mitigate supply chain disruptions, and provide critical capacity in military, public health preparedness, humanitarian, and emergency response contexts. Looking further into the future, the potential of centralized production opens opportunities for the bedside production of personalized medicine and perhaps even astropharmacy to support long-duration space flight^86^. The MANGO device itself was built to make assembly simple and accessible, with fluidic and electronic complexity simplified by embedding them into the manifold and PCB/Raspberry Pi, respectively. The design is low-cost and user-friendly, with open-source files available on GitHub for users and community-driven improvements. At approximately $1,500 USD, it is an estimated four times cheaper than the earlier BioMOD concept ($6,000 USD) and about half the size^48^.

The MANGO device is not without its limitations. As with any protein purification process, protocol development is required to tailor CFPS expression conditions^39, 86–91^ and fluidic volumes. This is achieved through manual control via the user interface **(Fig. 2a, b)** of each automated step (i.e., aeration during synthesis, purification, and elution), which was the primary modality for demonstrations here. However, as shown in Figure S19, once protocols are established, complete automation of expression and purification can be achieved for routine production. Our next steps will involve adding optical sensors throughout the device to improve operational efficiency and the collection of the eluted product. Further, while current 1 mL reactions can yield 0.1–0.5 mg of purified protein, sufficient for thousands of RT-LAMP tests, future designs could enable larger CFPS volumes to meet the needs of applications such as antibody or vaccine production.

We also wish to share some of the challenges that we faced in our efforts to validate MANGO abroad. While we have team members in South America, shipping devices and molecular components far from home brought challenges. These included customs delays, damaged, lost, or stolen packages requiring reshipment, and unexpected damage to sensitive components needing on-site troubleshooting and repair. We have previously shared tips on how to deploy efforts to remote sites from your home lab^8, 13^. While certainly challenging, we find that real-world testing provides valuable feedback on designs and offers opportunities to work with a team of practical users who are familiar with local needs and challenges.

Importantly, MANGO was built and field-validated for research use only (RUO). According to current FDA guidance^87^, RUO reagents are not required to be manufactured under current Good Manufacturing Practices (GMPs), which has enabled us to develop a framework for rapid and cost-effective bioreagent production on MANGO. Looking forward, the open-source architecture and modular control framework of MANGO lay a foundation for compatibility with GMP-compliant workflows as the technology advances toward clinical or diagnostic applications. Additionally, MANGO produced enzymes that matched commercial gold standard reagents in human sample testing **(Figs. 5 and 6)**, demonstrating that decentralized production can supply high-quality products for developing and validating new *in vitro* diagnostic tests Across well-resourced and resource-limited settings, we see MANGO as a catalyst that enables local biomanufacturing to add to existing GMP-grade products, ensuring essential bioreagents are available when needed with production costs adjusted to local economies and needs.

Advances in on-site DNA synthesis^88, 89^, along with compact, automated devices like MANGO, offer exciting glimpses of what the future may hold for research and healthcare. Much like a 3D printer, decentralized protein biomanufacturing has far-reaching implications, where any coding sequence can be turned into a functional protein using only basic reagents and a digital file. This integration could dramatically expand access to biologics and serve as a strategic biotech capacity for military deployments and autonomous health and research operations in space as humanity pushes beyond low Earth orbit. More broadly, decentralizing science and engineering can drive community-driven solutions, strengthen connections, enhance global capacity, and ensure that the benefits of biotechnology are shared equitably. By empowering communities to define problems, ask research questions, and own the tools to solve them, bioengineering fosters innovation where it matters most.

## METHODS

### Bacterial strains

*E. coli* BL21 (DE3) (NEB, C2527I), *E. coli* BL21 Star (DE3) (Invitrogen, C601003), ClearColi BL21(DE3) (Biosearch Technologies, 60810-1), and *E. coli* SHuffle (NEB, C3028J) strains were used to prepare cell-free lysates. *E. coli* NEB 5-alpha (NEB, C2987I) was employed for plasmid cloning and purification. Bacterial strains were cultured under standard conditions (LB medium, 37 °C, shaking incubator), except when grown for the preparation of cell-free lysates.

### Mammalian cell culture

Fibroblast L-929 cells (ATCC; CCL-1) were cultured in DMEM supplemented with 10% FBS and maintained under standard culture conditions (37 °C, 5% CO₂). These cultures served as the cellular system for subsequent TNF neutralization assays.

### Virus culture and propagation

Patient-derived strains of DENV-2 (PE/95-3808) and SARS-CoV-2 (46519/Brazil/PE-FIOCRUZ-IAM4372/2021) were isolated in Brazil and subsequently propagated in Vero and Vero-E6 cells, respectively. Viral titers, measured by the 50% Tissue Culture Infectious Dose (TCID_50_) assay, ranged from 10^6^ to 10^7^ TCID_50_/mL. Aliquots were prepared and stored at –80 °C as viral stocks for use in downstream assays.

### DNA template design and preparation

DNA sequences encoding the proteins of interest were obtained from the scientific literature, patents, or deposited Addgene constructs. They were codon-optimized for expression in *E. coli* K-12 using the Integrated DNA Technologies (IDT) codon optimization web tool. When necessary, ribosome binding strength was assessed with the RBS calculator^90^, and existing or predicted 3D structures were analyzed to guide tag placement. To facilitate downstream affinity purification, constructs were engineered to include either a His₆ or Strep-tag II at the N- or C-terminus. Cloning and plasmid propagation were conducted using NEBuilder HiFi DNA Assembly Master Mix (NEB, E2621) and competent *E. coli* cells (NEB, C2987I). For restriction enzyme production, synthetic DNA fragments corresponding to the target genes were obtained as linear constructs containing Ter sites^91^ (Twist Bioscience), which were PCR-amplified before use as templates for protein expression. For RT-LAMP enzyme manufacturing and nanobody development, synthetic DNA fragments encoding the target protein and required tags were ordered from Twist Bioscience and cloned into the T7p14 expression plasmid. Alternatively, these fragments were ordered as cloned genes in T7p14 from Twist Bioscience. **All DNA sequences are provided in Supplementary Data 1 and 2.**

### In-house cell-free extract preparation

Cell-free lysate was prepared from *E. coli* strains^92^, unless otherwise stated. Briefly, 1 L of culture was prepared in 4 L baffled flasks using 2x Yeast Tryptone Peptone Glucose (YTPG) medium [molecular-grade water supplemented with 20% (w/v) yeast extract, 32% (w/v) tryptone, 10% (w/v) sodium chloride (NaCl), 44 mM potassium phosphate monobasic (KH₂PO₄), 80 mM potassium phosphate dibasic (K₂HPO₄), and 100 mM glucose, pH 7.2], and incubated at 37 °C with shaking at 200 rpm. Cultures were induced with 1 mM isopropyl β-D-1-thiogalactopyranoside (IPTG) at OD₆₀₀ = 0.6 and harvested at OD₆₀₀ = 3.0. Cell pellets were washed three times with S30 buffer [10 mM Tris-acetate (pH 8.2), 14 mM magnesium acetate, 60 mM potassium acetate, supplemented with 2 mM dithiothreitol (DTT) immediately before use], using 25 mL per wash per pellet. Pellets were resuspended in 1 mL S30 buffer per gram of wet cells and lysed by sonication in an ice-water bath. Total sonication energy was calculated using the formula: Energy (J) = (Volume (µL) − 33.6)/1.8. A QSonica CL-18 (20 kHz) probe was applied at 50% amplitude with a cycle of 10 s on and 15 s off until the required energy was delivered. Following lysis, an additional 2 mM DTT was added, and cell lysates were clarified by centrifugation at 18,000 × g for 10 min at 4 °C. The supernatant was collected, incubated at 37 °C and 250 rpm for 1 hour, and then centrifuged again at 10,000 × g for 10 minutes at 4 °C. The resulting supernatant was collected, flash-frozen, and stored at −80 °C.

### General cell-free reaction preparation

Cell-free reactions followed the protocol previously described^92^ with the following modifications: 3-phosphoglyceric acid (3-PGA) was used in place of phosphoenolpyruvate, putrescine was omitted, and PEG 8k was added at 2% (w/v). The amino acid mixture was prepared as described previously^93^. Final concentrations in the cell-free reactions were: 1.2 mM ATP, 0.85 mM CTP, 0.85 mM UTP, 0.85 mM GTP, 31.5 μg/mL folinic acid, 170.6 μg/mL tRNA, 0.40 mM NAD, 0.27 mM CoA, 4.0 mM oxalic acid, 1.5 mM spermidine, 57.3 mM HEPES buffer, 3 mM of each amino acid, and 30 mM 3-PGA. Salt components were typically added as 10 mM magnesium glutamate and 130 mM potassium glutamate, although concentrations were adjusted when necessary. DNA was included at a final concentration of 15 nM, and crude cell lysate constituted 33.3% of the total reaction volume. Following assembly, CFPS reactions were loaded into the MANGO device and incubated overnight at room temperature, enabling protein expression and subsequent purification. CFPS reactions using commercial cell-free lysates (Liberum, CF0001.B) were prepared according to the manufacturer’s instructions. To remove insoluble protein fractions and aggregates, the CFPS reactions were centrifuged at 20,000 × g for 5 minutes, and the resulting clarified supernatant was collected for protein purification. Protein quantification was performed with the Pierce BCA protein assay kit (Thermo Fisher, 23225). All purification fractions were analyzed by SDS-PAGE, and band signal intensities were quantified using ImageJ software (v1.53k).

### Lyophilization of molecular components

CFPS reactions were lyophilized as previously described^8^. The lyophilized samples were then packaged into vacuum-sealable bags with three desiccant packs and two oxygen absorbers, flushed with nitrogen gas again, and sealed with an impulse heat vacuum sealer. FD-CFPS reactions were stored either at room temperature on the lab bench or shipped overseas without cold chain logistics, reaching collaborators in Brazil and Colombia.

### MANGO fabrication

Briefly, the main body, control unit housing, and holders for the fluidic manifold and driver board were fabricated using 3D printing on an Ultimaker 3 Extended (Ultimaker, Netherlands) with 2.85-mm PLA filament. The enclosure for the MANGO device was custom-designed by BioHues Digital (https://biohuesdigital.com/). Smooth manifold movement was enabled by stainless steel rods fitted with 6-mm linear ball bearings. Mounting points for the IncuKit MINI thermostat, fan, and heater (Incubator Warehouse) were 3D-printed and secured to the enclosure with screws. For reliable closure, neodymium magnets (1 cm diameter) were positioned at each corner of the door interface. The fluidic manifold was fabricated by precision laser-cutting flow channels into a 140-µm-thick ARcare® 90106NB double-sided adhesive spacer (Adhesive Research). Pump and valve mounts, along with ports for the purification column and buffer/reaction tubes, were CNC-milled into two 6.35-mm-thick cast acrylic plates (McMaster-Carr). These plates were bonded with the patterned adhesive under overnight pressure, after which miniature pumps, valves (The Lee Company), and fluid connectors were installed. The assembled manifold was secured to its 3D-printed holder with stainless steel screws and nuts. MANGO fabrication details are available on GitHub and can be modified as needed.

Purification columns were prepared by inserting a 3-mm punched filter (Pierce™ Spin Columns, Thermo Fisher) into 7-cm FEP tubing (ID 1/8”, OD 3/16“; GE Healthcare) with barbed fittings (McMaster-Carr). **See the Supplementary Information for the detailed protocol.**

The control unit included a Raspberry Pi 4 (4 GB RAM, 128 GB SD card; Raspberry Pi Foundation) with a screw-terminal breakout (GeeekPi) mounted on a 7-inch LCD touchscreen. A 5A DC–DC buck converter stepped down the 12V input power to the 5V required by the Raspberry Pi, and two DC cooling fans were installed in the enclosure for thermal management. The Raspberry Pi was connected to the driver board in the main unit via a multi-pin circular connector. Each pump and valve is connected to its corresponding terminal on the driver board.

### MANGO operation

MANGO operates from AC or portable power. Upon startup, the incubator warms to a user-set temperature, and a Python-based control software launches on the integrated touchscreen, allowing for the programming of aeration frequency and CFPS duration (typically 16 hours). FD-CFPS reactions are rehydrated and mounted on the manifold, with aeration provided by a pulsatile pump (1 pulse s⁻¹, user-adjustable).

Purification buffers were prepared as described in detail **in the Supplementary Information**. Before purification, all fluidic lines were primed with 1.5 mL binding buffer. The purification column was equilibrated with 40 column volumes of binding buffer (1 pump s⁻¹), after which the CFPS mixture was loaded (1 pump s⁻¹) and residual protein flushed with 2 mL binding buffer (2 pumps s⁻¹). The column was then washed with 80 column volumes of wash buffer (2 pumps s⁻¹). Elution proceeded at a flow rate of 1 pump every 2 seconds. The first 0.6 mL of eluate, corresponding to column dead volume, was discarded. Following purification, the column was cleaned sequentially with 2 mL each of elution buffer, Milli-Q water, and 30% ethanol, wrapped in parafilm, and stored at 4 °C. Fluidic paths were cleaned by washing with 1 mL of 0.5 M NaOH followed by 2 mL of binding buffer, leaving the channels empty for subsequent use.

### Endotoxin removal and quantification

Endotoxin removal was performed using the Pierce High-Capacity Endotoxin Removal Spin Column Kit (Thermo Fisher Scientific, 88274), except when using cell-free lysates from Clear Coli. The endotoxin levels were then measured with the Pierce Chromogenic Endotoxin Quantification Kit (Thermo Fisher Scientific, A39552). Throughout all procedures, endotoxin-free ultrapure water (Sigma-Aldrich, TMS-011-A) was used.

### Expression, in situ biotinylation, and purification of deGFP

A construct encoding deGFP with an N-terminal His₆ tag and a C-terminal AVI tag was expressed in *E. coli* BL21(DE3) lysate (1 mL, 16 h, room temperature, aeration at 1 pulse s⁻¹) in the presence of BirA biotin–protein ligase (Avidity) to enable co-translational biotinylation. **See the Supplementary Information for the detailed protocol.**

### Nb21 dot blot assay

Dot blotting was performed to confirm the expression of the target protein as previously described^94^. For each sample, 0.5 μL of the CFPS reaction was applied to a nitrocellulose membrane and allowed to air dry. Membranes were blocked for 1 hour with 5% skim milk (Bioshop) in TBST, then incubated for 1 hour with anti-His₆-HRP Ab (SouthernBiotech, 4603-05, 1:8,000 dilution) in 0.05% milk in TBST. After five TBST washes (each lasting 5 minutes), the membranes were developed using Clarity Max ECL substrate (Bio-Rad) and imaged with the colorimetric and chemiluminescent settings of the Bio-Rad ChemiDoc MP Imaging System.

### Nb21 ELISA assay

ELISA was used to confirm that cell-free–produced nanobodies bind to a commercial SARS-CoV-2 spike antigen. Black 384-well plates were coated overnight at 4 °C with RBD antigen (SinoBiological [40592-VNAH], 1 μg/mL in PBS). Plates were then blocked for 1 hour at 350 rpm with 5% skim milk (Bioshop) in PBS (Gibco). Wells were washed six times with PBST (0.05% Tween-20 in PBS) and incubated for 1 h at 350 rpm with nanobodies (7.5 μg/mL) diluted in 0.5% skim milk (Bioshop) in PBS (Gibco). After six additional washes with PBST, wells were incubated for 1 hour at 350 rpm with anti-His₆-HRP Ab (SouthernBiotech, 4603-05, 1:6,000) prepared in 0.5% skim milk (Bioshop) in PBST (Gibco). Following six additional washes with PBST, detection was performed using the QuantaBlu fluorogenic peroxidase substrate kit (Thermo Fisher, 15169) according to the manufacturer’s instructions. Fluorescence was measured at 325 and 425 nm using a BioTek Neo microplate reader.

### TNF Nb characterization

TNF neutralization assays were performed using fibroblast L-929 cells. Cells were seeded at ∼25% confluency and cultured overnight at 37 °C with 5% CO₂. On the following day, the medium was replaced with fresh medium that had been pre-incubated (1 h, 37 °C, 5% CO₂) with antibodies (133 nM) and TNF (≤100 ng/mL). The assay included anti-TNF Nb (MANGO), commercial anti-TNF IgG (SinoBiological, 50349-RN023-2), and commercial TNF (SinoBiological, 50349-MNAE), with Nb21 (MANGO) serving as a negative control. Actinomycin D was then added to each well (final concentration 1 µM), and cells were incubated overnight. Cell viability was assessed by adding CellTiter-Glo 2.0 reagent (Promega, 9242) at a 1:1 ratio with culture medium. Plates were shaken at 300 rpm for 2 min, and luminescence was recorded. Cells treated with 1% Triton X-100 (0% survival) and cells not exposed to TNF (100% survival) served as lower and upper normalization controls, respectively.

### Restriction endonuclease activity assay

The catalytic activity of MANGO-expressed restriction endonucleases benchmarked against their commercial counterparts from NEB (NcoI-HF, R3193S; EcoRI-HF, R3101S; HindIII-HF, R3104S). Briefly, 50 µL reactions were prepared with 1X rCutSmart Buffer (NEB, B6004S), 1 µg of a 1 kb linear DNA substrate containing the relevant restriction site near the center, and either 20 U EcoRI or 10 U HindIII/NcoI. Reactions were incubated at 37 °C for 30 minutes. After incubation, samples were separated by electrophoresis on 2% agarose gels. **See the Supplementary Information for detailed protocols and optimization experiments.**

### T4 DNA ligase–mediated DNA joining assay

To evaluate the activity of MANGO-expressed T4 DNA ligase, a 1 kb DNA fragment was first digested with HindIII, EcoRI, or NcoI derived from MANGO, following the digestion protocol described above. Following restriction digestion and enzyme inactivation, ligation reactions were assembled by adding 1X T4 ligase buffer (Promega, C1263) and 500 nM MANGO-derived T4 DNA ligase. Reactions were incubated for 1 h at 30 °C, then analyzed by agarose gel electrophoresis. **See the Supplementary Information for detailed protocols and optimization experiments.**

### Mosquito sample testing under controlled conditions

To assess the performance of the RT-LAMP assay for detecting DENV-2 in *Aedes aegypti* mosquito samples, crude lysates were prepared from mosquito pools (10 mosquitoes per pool). The lysates were then spiked with cultured DENV-2 to achieve final viral loads of 10⁶ (high) or 10³ (low) PFU/mL, as previously reported^60^, and directly processed and tested by RT-LAMP.

### Synthetic RNA controls for molecular reactions

Synthetic positive controls were obtained as gene fragments from Twist Bioscience or IDT **(Supplementary Data 3 and 4)**. Viral targets were first PCR-amplified using Q5 High-Fidelity DNA Polymerase (NEB, M0491L). To produce RNA targets, a T7 promoter sequence was introduced during PCR, and the resulting linear DNA was subsequently transcribed *in vitro* with the HiScribe T7 Quick High Yield RNA Synthesis Kit (NEB, E2050S) **(Supplementary Data 5)**. Synthetic RNA controls were stored at -80°C until they were used in downstream applications. A synthetic SARS-CoV-2 RNA was acquired from Twist Bioscience (102024) and used as a positive control in SARS-CoV-2 detection assays.

### Nucleic acid isolation

Viral RNA was isolated from cultured viruses or clinical samples, such as nasopharyngeal swabs or serum, using the QIAamp Viral RNA Mini Kit (Qiagen, 52904) according to the manufacturer’s instructions. The processed RNA was stored at –80 °C and subsequently used as input for molecular reactions.

### RT-qPCR assay for SARS-CoV-2 and DENV detection

Nasopharyngeal swab or serum samples were tested for positivity and analyzed using RT-qPCR, following protocols established by the U.S. Centers for Disease Control and Prevention (CDC). RT-qPCR reactions were performed with the QuantiNova Probe RT-PCR Kit (Qiagen, 208354) according to the manufacturer’s instructions. In brief, each reaction was prepared using the QuantiNova Probe RT-PCR kit according to the manufacturer’s protocols, with a final volume of 10 μL. RT-qPCR reactions were performed in 96- or 384-well plates using primers at a final concentration of 800 nM and probes at 100 nM, with 3.5 µL of template as the input for each reaction. All reactions were conducted using the Applied Biosystems QuantStudio 3 or 5 Real-Time PCR Systems (Applied Biosystems, USA). All oligos were synthesized by IDT and **are listed in Supplementary Table 4.**

### RT-LAMP reaction setup

RT-LAMP reactions (10 µL) were prepared with 1X isothermal buffer (20 mM Tris-HCl, 10 mM (NH₄)₂SO₄, 50 mM KCl, 2 mM MgSO₄, 0.1% Tween-20, pH 8.8), 7.31 ng/µL Bst DNA Polymerase LF (produced on MANGO), and 2.15 ng/µL M-MLV Reverse Transcriptase (produced on MANGO), 4 mM MgSO₄, 1.4 mM dNTPs (NEB, N0446S), and primers at the following concentrations: 0.2 μM for F3 and B3, 1.6 μM for FIP and BIP, 0.4 μM for LF and LB. Each reaction contained 1.0 μL of template (in vitro transcribed RNA or RNA isolated from patient samples) or nuclease-free water for non-template control (NTC) reactions. In-house isothermal and salt buffers were prepared according to protocols available online (https://www.protocols.io/view/lamp-rt-lamp-buffer-protocol-j8nlk4351g5r/v1). For benchmarking, 10 μL reactions were also prepared using WarmStart LAMP Master Mix (NEB, E1700S) following the manufacturer’s instructions. Reactions were assembled on ice and incubated at 63–65 °C for 20–40 minutes using a qPCR machine, a conventional thermal cycler, or FluoroPLUM (LSK Technologies, now part of Nicoya), depending on the target-specific conditions, and then heat-inactivated at 80 °C for 5 minutes.

RT-LAMP reactions were analyzed as described previously, using either fluorescent, visual colorimetric readouts, or gel electrophoresis^60, 63^. Fluorescent dyes, including LAMP dye (1X final concentration, NEB, B1700S) and SYTO-9 (10 μM final concentration, Invitrogen, S34854)^60, 63^, were used for real-time fluorescence monitoring, while SYBR Gold Nucleic Acid Stain (diluted 1:10 in nuclease-free water, Invitrogen, S11494) was used for endpoint visual colorimetric detection^60^. For colorimetric detection, reaction tubes were photographed with a smartphone, and the amplicons were analyzed through electrophoresis on 1.5% agarose gels. All oligonucleotides were synthesized by IDT and **are listed in Supplementary Data 3.**

### Data analysis and statistics

All experiments were performed independently at least three times, with three technical replicates each. Graphs and statistical analyses (t-test and ANOVA) were performed using GraphPad Prism (version 10.6.0), unless otherwise noted. Diagnostic performance analyses were performed using MedCalc’s online mathematical software (https://www.medcalc.org/calc/diagnostic_test.php).

### Patient sample collection

Clinical specimens were obtained in Canada, Brazil, and Colombia. In Canada, nasopharyngeal swabs were collected from patients with suspected respiratory infections at the Mount Sinai Hospital diagnostic laboratory in Toronto. In Brazil, nasopharyngeal and serum samples were obtained through the Fiocruz Pernambuco genomic surveillance network and later provided for this study. In Colombia, serum samples were collected from patients diagnosed with DENV using a rapid test at the Hospital Universitario Hernando Moncaleano Perdomo in Neiva, Huila, and then supplied for this study. Additional serum samples from patients exhibiting febrile symptoms suggestive of arboviral infection were transferred to CIMPAT under a Material Transfer Agreement (MTA) for investigation purposes. In accordance with quality control, only patient samples that tested positive for the endogenous human control gene were included in the study. **Ethics**

This study was approved by the Research Ethics Board (REB) at the University of Toronto (protocol #46252 for arbovirus studies and #39531 for COVID-19 studies), by the Institutional Review Board of HEMOPE-PE, Brazil (CAAE: 43877521.4.00000.5195), and by the Ethics Committee of Hospital Universitario Hernando Moncaleano Perdomo, Colombia (protocol #02-07). All clinical trials were carried out in accordance with local and international regulations, including the World Medical Association’s Declaration of Helsinki.

## Data availability

Source data supporting the findings of this study are included within this paper and the Supplementary Information. Detailed instructions for building the MANGO device are available on our GitHub repository, ensuring open access and reproducibility. DNA sequence information for the constructs and oligos used in this study can be found in Supplementary Data.

## Acknowledgements

We thank all members of the participating laboratories in this international, multi-site research effort for their valuable technical support, insightful discussions, and critical feedback. We also thank Mayara Matias de Oliveira Marques da Costa, Gustavo Barbosa de Lima, Andreza Pâmela Vasconcelos, Keilla Maria Paz e Silva, Diego Arruda Falcão, and other healthcare professionals for their essential role in processing patient samples from Brazil and Canada. Additionally, we thank the Secretaría de Salud del Putumayo (collaboration agreement #306-2024 with Uniandes) for their assistance in processing patient samples from Colombia. We are grateful to Dr. Constância Ayres, Dr. Larissa Krokovsky, and Dr. Duschinka Guedes from the Department of Entomology at the Oswaldo Cruz Foundation (Fiocruz) for generously providing the mosquitoes used in this study. We also extend our sincere appreciation to the staff at all participating institutions for their exceptional support in hosting and assisting visiting researchers throughout the project.

## Author contributions

S.J.R.d.S., M.S., and Q.M. contributed equally. S.J.R.d.S., M.S., Q.M., I.A.I., and S.S. co-wrote the manuscript. S.J.R.d.S., M.S., Q.M., J.R.J.V., S.S., I.A.I., B.N.R.S., D.D., P.B.B., K.B., and L.A.C. designed and carried out most of the experimental work. K.P. and D.S. supervised the development and fabrication of the MANGO device across multiple generations. Contributions to early versions were made by F.M. (generations 1-3), S.T. (generation 1), P.B. (generations 2-3), and R.F., W.H., and M.S. (generation 3). M.N. designed the purification column. M.S. designed the electronics and software for the final system, performed its assembly and characterization, developed the operating protocols, and conducted all protein purifications in Toronto. Q.M., M.S., and K.B. established the cell-free expression and purification of deGFP. Q.M., M.S., and L.A.C. established protocols for nanobodies. Q.M., M.S., and I.A.I. established protocols for restriction enzymes. S.J.R.d.S., M.S., and Q.M. established protocols for Bst LF polymerase and M-MLV reverse transcriptase. S.J.R.d.S., M.S., Q.M., J.R.J.V., A.B., C.G., L.P., and K.P. coordinated the deployment and field validation of MANGO in South America. S.J.R.d.S. built the RT-LAMP and RT-qPCR assays. S.J.R.d.S. developed and implemented the diagnostic testing pipeline for pathogen detection through clinical trials in Canada, Brazil, and Colombia. Y.G., S.C., and K.P. developed the FluoroPLUM. S.S. and L.A.C. performed the nanobody experiments. I.A.I. performed experiments with the cloning enzymes. S.J.R.d.S., J.J.F.d.M., C.F.N., M.H.S.P., G.L.W., T.M., C.G., and L.P. contributed to the collection and acquisition of patient samples. S.J.R.d.S., Q.M., J.R.J.V., B.N.R.S., D.D., P.B.B., R.P.G.M., C.L., and L.C.M. contributed to the clinical trials. A.R., M.C., and A.B. supported the project. S.J.R.d.S., M.S., Q.M., P.B., J.R.J.V., S.S., I.A.I., B.N.R.S., K.B., and L.A.C. performed data analysis, statistics, and interpreted results. K.P. supervised the project, designed experiments, and co-wrote the manuscript with L.P. and C.G. All authors edited and approved the final version of the manuscript.

## REFERENCES

(1) Overton, T. W. Recombinant protein production in bacterial hosts. Drug Discov Today 2014, 19 (5), 590–601. DOI: 10.1016/j.drudis.2013.11.008.

(2) Pardee, K.; Slomovic, S.; Nguyen, P. Q.; Lee, J. W.; Donghia, N.; Burrill, D.; Ferrante, T.; McSorley, F. R.; Furuta, Y.; Vernet, A.;, et al. Portable, On-Demand Biomolecular Manufacturing. Cell 2016, 167 (1), 248–259.e212. DOI: 10.1016/j.cell.2016.09.013.

(3) Kumraj, G.; Pathak, S.; Shah, S.; Majumder, P.; Jain, J.; Bhati, D.; Hanif, S.; Mukherjee, S.; Ahmed, S. Capacity Building for Vaccine Manufacturing Across Developing Countries: The Way Forward. Hum Vaccin Immunother 2022, 18 (1), 2020529. DOI: 10.1080/21645515.2021.2020529.

(4) Lee, S. J.; Kim, D. M. Cell-Free Synthesis: Expediting Biomanufacturing of Chemical and Biological Molecules. Molecules 2024, 29 (8). DOI: 10.3390/molecules29081878.

(5) Watson, J. L.; Juergens, D.; Bennett, N. R.; Trippe, B. L.; Yim, J.; Eisenach, H. E.; Ahern, W.; Borst, A. J.; Ragotte, R. J.; Milles, L. F.;, et al. De novo design of protein structure and function with RFdiffusion. Nature 2023, 620 (7976), 1089–1100. DOI: 10.1038/s41586-023-06415-8.

(6) Tsuboyama, K.; Dauparas, J.; Chen, J.; Laine, E.; Mohseni Behbahani, Y.; Weinstein, J. J.; Mangan, N. M.; Ovchinnikov, S.; Rocklin, G. J. Mega-scale experimental analysis of protein folding stability in biology and design. Nature 2023, 620 (7973), 434–444. DOI: 10.1038/s41586-023-06328-6.

(7) Perez, J. G.; Stark, J. C.; Jewett, M. C. Cell-Free Synthetic Biology: Engineering Beyond the Cell. Cold Spring Harb Perspect Biol 2016, 8 (12). DOI: 10.1101/cshperspect.a023853.

(8) Silva, S. J.; al, e. International Multi-site Implementation of Local Cell-Free Protein Biomanufacturing to Advance Health and Research Equity. In medRxiv, 2025.

(9) Matthews, Q.; da Silva, S. J. R.; Norouzi, M.; Pena, L. J.; Pardee, K. Adaptive, diverse and de-centralized diagnostics are key to the future of outbreak response. BMC Biol 2020, 18 (1), 153. DOI: 10.1186/s12915-020-00891-4.

(10) Duma, Z.; Chuturgoon, A. A.; Ramsuran, V.; Edward, V.; Naidoo, P.; Mpaka-Mbatha, M. N.; Bhengu, K. N.; Nembe, N.; Pillay, R.; Singh, R.;, et al. The challenges of severe acute respiratory syndrome coronavirus 2 (SARS-CoV-2) testing in low-middle income countries and possible cost-effective measures in resource-limited settings. Global Health 2022, 18 (1), 5. DOI: 10.1186/s12992-022-00796-7.

(11) Cornish, N. E.; Bachmann, L. H.; Diekema, D. J.; McDonald, L. C.; McNult, P.; Stevens-Garcia, J.; Raphael, B. H.; Miller, M. B. Pandemic Demand for SARS-CoV-2 Testing Led to Critical Supply and Workforce Shortages in U.S. Clinical and Public Health Laboratories. J Clin Microbiol 2023, 61 (7), e0318920. DOI: 10.1128/jcm.03189-20.

(12) Silva, S. J. R. D.; Magalhães, J. J. F.; Pena, L. Simultaneous Circulation of DENV, CHIKV, ZIKV and SARS-CoV-2 in Brazil: an Inconvenient Truth. One Health 2021, 12, 100205. DOI: 10.1016/j.onehlt.2020.100205.

(13) Karlikow, M.; da Silva, S. J. R.; Guo, Y.; Cicek, S.; Krokovsky, L.; Homme, P.; Xiong, Y.; Xu, T.; Calderón-Peláez, M. A.; Camacho-Ortega, S.;, et al. Field validation of the performance of paper-based tests for the detection of the Zika and chikungunya viruses in serum samples. Nat Biomed Eng 2022, 6 (3), 246–256. DOI: 10.1038/s41551-022-00850-0.

(14) da Silva, S. J. R.; do Nascimento, J. C. F.; Germano Mendes, R. P.; Guarines, K. M.; Targino Alves da Silva, C.; da Silva, P. G.; de Magalhães, J. J. F.; Vigar, J. R. J.; Silva-Júnior, A.; Kohl, A.;, et al. Two Years into the COVID-19 Pandemic: Lessons Learned. ACS Infect Dis 2022. DOI: 10.1021/acsinfecdis.2c00204.

(15) Wulff, R. T.; Qiu, Y.; Wu, C.; Calfee, D. P.; Singh, H. K.; Hatch, I.; Steel, P. A. D.; Scofi, J. E.; Westblade, L. F.; Cushing, M. M. Laboratory Interventions to Eliminate Unnecessary Rapid COVID-19 Testing During a Reagent Shortage. Am J Clin Pathol 2022, 158 (3), 401–408. DOI: 10.1093/ajcp/aqac063.

(16) Nouvellet, P.; Garske, T.; Mills, H. L.; Nedjati-Gilani, G.; Hinsley, W.; Blake, I. M.; Van Kerkhove, M. D.; Cori, A.; Dorigatti, I.; Jombart, T.;, et al. The role of rapid diagnostics in managing Ebola epidemics. Nature 2015, 528 (7580), S109–116. DOI: 10.1038/nature16041.

(17) Lippi, G.; Favaloro, E. J.; Plebani, M. Laboratory medicine and natural disasters: are we ready for the challenge? Clin Chem Lab Med 2010, 48 (5), 573–575. DOI: 10.1515/CCLM.2010.148.

(18) Bastani, P.; Sadeghkhani, O.; Bikine, P.; Mehralian, G.; Samadbeik, M.; Ravangard, R. Medication supply chain resilience during disasters: exploration of causes, strategies, and consequences applying Strauss and Corbin’s approach to the grounded theory. J Pharm Policy Pract 2023, 16 (1), 99. DOI: 10.1186/s40545-023-00604-6.

(19) Dahl, V.; Migliori, G. B.; Lange, C.; Wejse, C. War in Ukraine: an immense threat to the fight against tuberculosis. Eur Respir J 2022, 59 (4). DOI: 10.1183/13993003.00493-2022.

(20) Kearney, J. E.; Thiel, N.; El-Taher, A.; Akhter, S.; Townes, D. A.; Trehan, I.; Pottinger, P. S. Conflicts in Gaza and around the world create a perfect storm for infectious disease outbreaks. PLOS Glob Public Health 2024, 4 (2), e0002927. DOI: 10.1371/journal.pgph.0002927.

(21) Kluge, H. H. P.; Jakab, Z.; Bartovic, J.; D’Anna, V.; Severoni, S. Refugee and migrant health in the COVID-19 response. Lancet 2020, 395 (10232), 1237–1239. DOI: 10.1016/S0140-6736(20)30791-1.

(22) (CBD), C. o. B. D. Decision Adopted by the Conference of the Parties to the Convention on Biological Diversity: COP-16 Decision 21; 2024. www.cbd.int/doc/decisions/cop-16/cop-16-dec-21-en.pdf.

(23) WHO. Accelerating access to genomics for global health: promotion, implementation, collaboration, and ethical, legal, and social issues: a report of the WHO Science Council. 2022.

(24) Ortega, R. Scientists in Latin America struggle to get key chemicals and other reagents for experiments. A group has begun to help. In Science, 2024.

(25) Guzman-Chavez, F.; Arce, A.; Adhikari, A.; Vadhin, S.; Pedroza-Garcia, J. A.; Gandini, C.; Ajioka, J. W.; Molloy, J.; Sanchez-Nieto, S.; Varner, J. D.;, et al. Constructing Cell-Free Expression Systems for Low-Cost Access. ACS Synth Biol 2022, 11 (3), 1114–1128. DOI: 10.1021/acssynbio.1c00342.

(26) Jung, J. K.; Alam, K. K.; Verosloff, M. S.; Capdevila, D. A.; Desmau, M.; Clauer, P. R.; Lee, J. W.; Nguyen, P. Q.; Pastén, P. A.; Matiasek, S. J.;, et al. Cell-free biosensors for rapid detection of water contaminants. Nat Biotechnol 2020, 38 (12), 1451–1459. DOI: 10.1038/s41587-020-0571-7.

(27) Stark, J. C.; Jaroentomeechai, T.; Moeller, T. D.; Hershewe, J. M.; Warfel, K. F.; Moricz, B. S.; Martini, A. M.; Dubner, R. S.; Hsu, K. J.; Stevenson, T. C.;, et al. On-demand biomanufacturing of protective conjugate vaccines. Sci Adv 2021, 7 (6). DOI: 10.1126/sciadv.abe9444.

(28) Warfel, K. F.; Williams, A.; Wong, D. A.; Sobol, S. E.; Desai, P.; Li, J.; Chang, Y. F.; DeLisa, M. P.; Karim, A. S.; Jewett, M. C. A Low-Cost, Thermostable, Cell-Free Protein Synthesis Platform for On-Demand Production of Conjugate Vaccines. ACS Synth Biol 2023, 12 (1), 95–107. DOI: 10.1021/acssynbio.2c00392.

(29) Stark, J. C.; Huang, A.; Nguyen, P. Q.; Dubner, R. S.; Hsu, K. J.; Ferrante, T. C.; Anderson, M.; Kanapskyte, A.; Mucha, Q.; Packett, J. S.;, et al. BioBits™ Bright: A fluorescent synthetic biology education kit. Sci Adv 2018, 4 (8), eaat5107. DOI: 10.1126/sciadv.aat5107.

(30) Hunt, J. P.; Zhao, E. L.; Soltani, M.; Frei, M.; Nelson, J. A. D.; Bundy, B. C. Streamlining the preparation of “endotoxin-free” ClearColi cell extract with autoinduction media for cell-free protein synthesis of the therapeutic protein crisantaspase. Synth Syst Biotechnol 2019, 4 (4), 220–224. DOI: 10.1016/j.synbio.2019.11.003.

(31) Salehi, A. S.; Smith, M. T.; Bennett, A. M.; Williams, J. B.; Pitt, W. G.; Bundy, B. C. Cell-free protein synthesis of a cytotoxic cancer therapeutic: Onconase production and a just-add-water cell-free system. Biotechnol J 2016, 11 (2), 274–281. DOI: 10.1002/biot.201500237.

(32) Welsh, J. P.; Lu, Y.; He, X. S.; Greenberg, H. B.; Swartz, J. R. Cell-free production of trimeric influenza hemagglutinin head domain proteins as vaccine antigens. Biotechnol Bioeng 2012, 109 (12), 2962–2969. DOI: 10.1002/bit.24581.

(33) Kanter, G.; Yang, J.; Voloshin, A.; Levy, S.; Swartz, J. R.; Levy, R. Cell-free production of scFv fusion proteins: an efficient approach for personalized lymphoma vaccines. Blood 2007, 109 (8), 3393–3399. DOI: 10.1182/blood-2006-07-030593.

(34) Sullivan, C. J.; Pendleton, E. D.; Sasmor, H. H.; Hicks, W. L.; Farnum, J. B.; Muto, M.; Amendt, E. M.; Schoborg, J. A.; Martin, R. W.; Clark, L. G.;, et al. A cell-free expression and purification process for rapid production of protein biologics. Biotechnol J 2016, 11 (2), 238–248. DOI: 10.1002/biot.201500214.

(35) Huang, A.; Nguyen, P. Q.; Stark, J. C.; Takahashi, M. K.; Donghia, N.; Ferrante, T.; Dy, A. J.; Hsu, K. J.; Dubner, R. S.; Pardee, K.;, et al. BioBits™ Explorer: A modular synthetic biology education kit. Sci Adv 2018, 4 (8), eaat5105. DOI: 10.1126/sciadv.aat5105.

(36) Smith, M. T.; Berkheimer, S. D.; Werner, C. J.; Bundy, B. C. Lyophilized Escherichia coli-based cell-free systems for robust, high-density, long-term storage. Biotechniques 2014, 56 (4), 186–193. DOI: 10.2144/000114158.

(37) Pardee, K.; Green, A. A.; Ferrante, T.; Cameron, D. E.; DaleyKeyser, A.; Yin, P.; Collins, J. J. Paper-based synthetic gene networks. Cell 2014, 159 (4), 940–954. DOI: 10.1016/j.cell.2014.10.004.

(38) Pardee, K.; Green, A. A.; Takahashi, M. K.; Braff, D.; Lambert, G.; Lee, J. W.; Ferrante, T.; Ma, D.; Donghia, N.; Fan, M.;, et al. Rapid, Low-Cost Detection of Zika Virus Using Programmable Biomolecular Components. Cell 2016, 165 (5), 1255–1266. DOI: 10.1016/j.cell.2016.04.059.

(39) Hunt, A. C.; Rasor, B. J.; Seki, K.; Ekas, H. M.; Warfel, K. F.; Karim, A. S.; Jewett, M. C. Cell-Free Gene Expression: Methods and Applications. Chem Rev 2025, 125 (1), 91–149. DOI: 10.1021/acs.chemrev.4c00116.

(40) Goerke, A. R.; Swartz, J. R. Development of cell-free protein synthesis platforms for disulfide bonded proteins. Biotechnol Bioeng 2008, 99 (2), 351–367. DOI: 10.1002/bit.21567.

(41) Buntru, M.; Vogel, S.; Stoff, K.; Spiegel, H.; Schillberg, S. A versatile coupled cell-free transcription-translation system based on tobacco BY-2 cell lysates. Biotechnol Bioeng 2015, 112 (5), 867–878. DOI: 10.1002/bit.25502.

(42) Kightlinger, W.; Duncker, K. E.; Ramesh, A.; Thames, A. H.; Natarajan, A.; Stark, J. C.; Yang, A.; Lin, L.; Mrksich, M.; DeLisa, M. P.;, et al. A cell-free biosynthesis platform for modular construction of protein glycosylation pathways. Nat Commun 2019, 10 (1), 5404. DOI: 10.1038/s41467-019-12024-9.

(43) Ramm, F.; Kaser, D.; König, I.; Fellendorf, J.; Wenzel, D.; Zemella, A.; Papatheodorou, P.; Barth, H.; Schmidt, H. Synthesis of biologically active Shiga toxins in cell-free systems. Sci Rep 2024, 14 (1), 6043. DOI: 10.1038/s41598-024-56190-3.

(44) Hellman, S.; Frisch, P.; Platzman, A.; Booth, P. 3D Printing in a hospital: Centralized clinical implementation and applications for comprehensive care. Digit Health 2023, 9, 20552076231221899. DOI: 10.1177/20552076231221899.

(45) Dodziuk, H. Applications of 3D printing in healthcare. Kardiochir Torakochirurgia Pol 2016, 13 (3), 283–293. DOI: 10.5114/kitp.2016.62625.

(46) Adamo, A.; Beingessner, R. L.; Behnam, M.; Chen, J.; Jamison, T. F.; Jensen, K. F.; Monbaliu, J. C.; Myerson, A. S.; Revalor, E. M.; Snead, D. R.;, et al. On-demand continuous-flow production of pharmaceuticals in a compact, reconfigurable system. Science 2016, 352 (6281), 61–67. DOI: 10.1126/science.aaf1337.

(47) Crowell, L. E.; Lu, A. E.; Love, K. R.; Stockdale, A.; Timmick, S. M.; Wu, D.; Wang, Y. A.; Doherty, W.; Bonnyman, A.; Vecchiarello, N.;, et al. On-demand manufacturing of clinical-quality biopharmaceuticals. Nat Biotechnol 2018. DOI: 10.1038/nbt.4262.

(48) Adiga, R.; Al-Adhami, M.; Andar, A.; Borhani, S.; Brown, S.; Burgenson, D.; Cooper, M. A.; Deldari, S.; Frey, D. D.; Ge, X.;, et al. Point-of-care production of therapeutic proteins of good-manufacturing-practice quality. Nat Biomed Eng 2018, 2 (9), 675–686. DOI: 10.1038/s41551-018-0259-1.

(49) Zawada, J. F.; Yin, G.; Steiner, A. R.; Yang, J.; Naresh, A.; Roy, S. M.; Gold, D. S.; Heinsohn, H. G.; Murray, C. J. Microscale to manufacturing scale-up of cell-free cytokine production--a new approach for shortening protein production development timelines. Biotechnol Bioeng 2011, 108 (7), 1570–1578. DOI: 10.1002/bit.23103.

(50) Alexander, E.; Leong, K. W. Discovery of nanobodies: a comprehensive review of their applications and potential over the past five years. J Nanobiotechnology 2024, 22 (1), 661. DOI: 10.1186/s12951-024-02900-y.

(51) Sun, D.; Sang, Z.; Kim, Y. J.; Xiang, Y.; Cohen, T.; Belford, A. K.; Huet, A.; Conway, J. F.; Sun, J.; Taylor, D. J.;, et al. Potent neutralizing nanobodies resist convergent circulating variants of SARS-CoV-2 by targeting diverse and conserved epitopes. Nat Commun 2021, 12 (1), 4676. DOI: 10.1038/s41467-021-24963-3.

(52) Xiang, Y.; Nambulli, S.; Xiao, Z.; Liu, H.; Sang, Z.; Duprex, W. P.; Schneidman-Duhovny, D.; Zhang, C.; Shi, Y. Versatile and multivalent nanobodies efficiently neutralize SARS-CoV-2. Science 2020, 370 (6523), 1479–1484. DOI: 10.1126/science.abe4747.

(53) FDA. Guideline on validation of the limulus amebocyte lysate test as an end-product endotoxin test for human and animal parental drugs, biological products, and medical devices. 1987; pp 1–30.

(54) Lobstein, J.; Emrich, C. A.; Jeans, C.; Faulkner, M.; Riggs, P.; Berkmen, M. SHuffle, a novel Escherichia coli protein expression strain capable of correctly folding disulfide bonded proteins in its cytoplasm. Microb Cell Fact 2012, 11, 56. DOI: 10.1186/1475-2859-11-56.

(55) Dhuriya, Y. K.; Sharma, D. Necroptosis: a regulated inflammatory mode of cell death. J Neuroinflammation 2018, 15 (1), 199. DOI: 10.1186/s12974-018-1235-0.

(56) Metcalfe, C.; Dougall, T.; Bird, C.; Rigsby, P.; Behr-Gross, M. E.; Wadhwa, M.; Study, P. O. T. The first World Health Organization International Standard for infliximab products: A step towards maintaining harmonized biological activity. MAbs 2019, 11 (1), 13–25. DOI: 10.1080/19420862.2018.1532766.

(57) Hoseini, S. S.; Sauer, M. G. Molecular cloning using polymerase chain reaction, an educational guide for cellular engineering. J Biol Eng 2015, 9, 2. DOI: 10.1186/1754-1611-9-2.

(58) Ishino, S.; Ishino, Y. DNA polymerases as useful reagents for biotechnology - the history of developmental research in the field. Front Microbiol 2014, 5, 465. DOI: 10.3389/fmicb.2014.00465.

(59) Kennedy, M. A.; Hosford, C. J.; Azumaya, C. M.; Luyten, Y. A.; Chen, M.; Morgan, R. D.; Stoddard, B. L. Structures, activity and mechanism of the Type IIS restriction endonuclease PaqCI. Nucleic Acids Res 2023, 51 (9), 4467–4487. DOI: 10.1093/nar/gkad228.

(60) Ribeiro da Silva, S. J.; Ferraz de Magalhães, J. J.; Matthews, Q.; Lot Divarzak, A. L.; Germano Mendes, R. P.; Rodrigues Santos, B. N.; Guerra de Albuquerque Cabral, D.; Bezerra da Silva, J.; Kohl, A.; Pardee, K.; et al. Development and field validation of an RT-LAMP assay for the rapid detection of chikungunya virus in patient and mosquito samples. Clin Microbiol Infect 2024. DOI: 10.1016/j.cmi.2024.03.004.

(61) da Silva, S. J. R.; Pardee, K.; Balasuriya, U. B. R.; Pena, L. Development and validation of a one-step reverse transcription loop-mediated isothermal amplification (RT-LAMP) for rapid detection of ZIKV in patient samples from Brazil. Sci Rep 2021, 11 (1), 4111. DOI: 10.1038/s41598-021-83371-1.

(62) Silva, S. J. R. D.; Paiva, M. H. S.; Guedes, D. R. D.; Krokovsky, L.; Melo, F. L.; Silva, M. A. L. D.; Silva, A. D.; Ayres, C. F. J.; Pena, L. J. Development and Validation of Reverse Transcription Loop-Mediated Isothermal Amplification (RT-LAMP) for Rapid Detection of ZIKV in Mosquito Samples from Brazil. Sci Rep 2019, 9 (1), 4494. DOI: 10.1038/s41598-019-40960-5.

(63) Lu, S.; Duplat, D.; Benitez-Bolivar, P.; León, C.; Villota, S. D.; Veloz-Villavicencio, E.; Arévalo, V.; Jaenes, K.; Guo, Y.; Cicek, S.;, et al. Multicenter international assessment of a SARS-CoV-2 RT-LAMP test for point of care clinical application. PLoS One 2022, 17 (5), e0268340. DOI: 10.1371/journal.pone.0268340.

(64) Matute, T.; Nuñez, I.; Rivera, M.; Reyes, J.; Blázquez-Sánchez, P.; Arce, A.; Brown, A. J.; Gandini, C.; Molloy, J.; Ramírez-Sarmiento, C. A.;, et al. Homebrew reagents for low-cost RT-LAMP. J Biomol Tech 2021, 32 (3), 114–120. DOI: 10.7171/jbt.21-3203-006.

(65) Lanciotti, R. S.; Kosoy, O. L.; Laven, J. J.; Velez, J. O.; Lambert, A. J.; Johnson, A. J.; Stanfield, S. M.; Duffy, M. R. Genetic and serologic properties of Zika virus associated with an epidemic, Yap State, Micronesia, 2007. Emerg Infect Dis 2008, 14 (8), 1232–1239. DOI: 10.3201/eid1408.080287.

(66) Lanciotti, R. S.; Kosoy, O. L.; Laven, J. J.; Panella, A. J.; Velez, J. O.; Lambert, A. J.; Campbell, G. L. Chikungunya virus in US travelers returning from India, 2006. Emerg Infect Dis 2007, 13 (5), 764–767. DOI: 10.3201/eid1305.070015.

(67) Silva, S. J. R. D.; Kohl, A.; Pena, L.; Pardee, K. Clinical and laboratory diagnosis of monkeypox (mpox): Current status and future directions. iScience 2023, 26 (6), 106759. DOI: 10.1016/j.isci.2023.106759.

(68) Notomi, T.; Okayama, H.; Masubuchi, H.; Yonekawa, T.; Watanabe, K.; Amino, N.; Hase, T. Loop-mediated isothermal amplification of DNA. Nucleic Acids Res 2000, 28 (12), E63.

(69) Notomi, T.; Mori, Y.; Tomita, N.; Kanda, H. Loop-mediated isothermal amplification (LAMP): principle, features, and future prospects. J Microbiol 2015, 53 (1), 1–5. DOI: 10.1007/s12275-015-4656-9.

(70) Silva, S. J. R. D.; Pardee, K.; Pena, L. Loop-Mediated Isothermal Amplification (LAMP) for the Diagnosis of Zika Virus: A Review. Viruses 2019, 12 (1). DOI: 10.3390/v12010019.

(71) Dao Thi, V. L.; Herbst, K.; Boerner, K.; Meurer, M.; Kremer, L. P.; Kirrmaier, D.; Freistaedter, A.; Papagiannidis, D.; Galmozzi, C.; Stanifer, M. L.;, et al. A colorimetric RT-LAMP assay and LAMP-sequencing for detecting SARS-CoV-2 RNA in clinical samples. Sci Transl Med 2020, 12 (556). DOI: 10.1126/scitranslmed.abc7075.

(72) CDC. Research use only 2019-Novel Coronavirus (2019-nCoV) Real-time RT-PCR primers and probes. In WHO, 2020.

(73) Aoki, M. N.; de Oliveira Coelho, B.; Góes, L. G. B.; Minoprio, P.; Durigon, E. L.; Morello, L. G.; Marchini, F. K.; Riediger, I. N.; do Carmo Debur, M.; Nakaya, H. I.; et al. Colorimetric RT-LAMP SARS-CoV-2 diagnostic sensitivity relies on color interpretation and viral load. Sci Rep 2021, 11 (1), 9026. DOI: 10.1038/s41598-021-88506-y.

(74) Magalhaes, T.; Braga, C.; Cordeiro, M. T.; Oliveira, A. L. S.; Castanha, P. M. S.; Maciel, A. P. R.; Amancio, N. M. L.; Gouveia, P. N.; Peixoto-da-Silva, V. J.; Peixoto, T. F. L.;, et al. Zika virus displacement by a chikungunya outbreak in Recife, Brazil. PLoS Negl Trop Dis 2017, 11 (11), e0006055. DOI: 10.1371/journal.pntd.0006055.

(75) Azevedo, E. A. N.; Silva, A. F. D.; Silva, V. G. D.; Machado, L. C.; de Lima, G. B.; Ishigami, B. I. M.; Silva, K. M. P. E.; Costa, M. M. O. M.; Falcão, D. A.; Vasconcelos, A. P.;, et al. Genomic and phenotypic characterization of the Oropouche virus strain implicated in the 2022-24 large-scale outbreak in Brazil. J Med Virol 2024, 96 (10), e70012. DOI: 10.1002/jmv.70012.

(76) Krokovsky, L.;, et al. Arbovirus Surveillance in Field-Collected Mosquitoes From Pernambuco-Brazil, During the Triple Dengue, Zika and Chikungunya Outbreak of 2015-2017. In Frontiers in Tropical Diseases, 2022.

(77) Ayllón, T.; Campos, R. M.; Brasil, P.; Morone, F. C.; Câmara, D. C. P.; Meira, G. L. S.; Tannich, E.; Yamamoto, K. A.; Carvalho, M. S.; Pedro, R. S.;, et al. Early Evidence for Zika Virus Circulation among Aedes aegypti Mosquitoes, Rio de Janeiro, Brazil. Emerg Infect Dis 2017, 23 (8), 1411–1412. DOI: 10.3201/eid2308.162007.

(78) Cevallos, V.; Ponce, P.; Waggoner, J. J.; Pinsky, B. A.; Coloma, J.; Quiroga, C.; Morales, D.; Cárdenas, M. J. Zika and Chikungunya virus detection in naturally infected Aedes aegypti in Ecuador. Acta Trop 2018, 177, 74–80. DOI: 10.1016/j.actatropica.2017.09.029.

(79) Souza-Neto, J. A.; Powell, J. R.; Bonizzoni, M. Aedes aegypti vector competence studies: A review. Infect Genet Evol 2019, 67, 191–209. DOI: 10.1016/j.meegid.2018.11.009.

(80) Weaver, S. C.; Reisen, W. K. Present and future arboviral threats. Antiviral Res 2010, 85 (2), 328–345. DOI: 10.1016/j.antiviral.2009.10.008.

(81) Nunes, P. C. G.; Daumas, R. P.; Sánchez-Arcila, J. C.; Nogueira, R. M. R.; Horta, M. A. P.; Dos Santos, F. B. 30 years of fatal dengue cases in Brazil: a review. BMC Public Health 2019, 19 (1), 329. DOI: 10.1186/s12889-019-6641-4.

(82) Rückert, C.; Weger-Lucarelli, J.; Garcia-Luna, S. M.; Young, M. C.; Byas, A. D.; Murrieta, R. A.; Fauver, J. R.; Ebel, G. D. Impact of simultaneous exposure to arboviruses on infection and transmission by Aedes aegypti mosquitoes. Nat Commun 2017, 8, 15412. DOI: 10.1038/ncomms15412.

(83) Santiago, G. A.; Vergne, E.; Quiles, Y.; Cosme, J.; Vazquez, J.; Medina, J. F.; Medina, F.; Colón, C.; Margolis, H.; Muñoz-Jordán, J. L. Analytical and clinical performance of the CDC real time RT-PCR assay for detection and typing of dengue virus. PLoS Negl Trop Dis 2013, 7 (7), e2311. DOI: 10.1371/journal.pntd.0002311.

(84) Grubaugh, N. D.; Torres-Hernández, D.; Murillo-Ortiz, M. A.; Dávalos, D. M.; Lopez, P.; Hurtado, I. C.; Breban, M. I.; Bourgikos, E.; Hill, V.; López-Medina, E. Dengue Outbreak Caused by Multiple Virus Serotypes and Lineages, Colombia, 2023-2024. Emerg Infect Dis 2024, 30 (11), 2391–2395. DOI: 10.3201/eid3011.241031.

(85) Rodríguez-Morales, A. J.; López-Medina, E.; Arboleda, I.; Cardona-Ospina, J. A.; Castellanos, J. E.; Faccini-Martínez, Á.; Gallagher, E.; Hanley, R.; Lopez, P.; Mattar, S.;, et al. The Epidemiological Impact of Dengue in Colombia: A Systematic Review. Am J Trop Med Hyg 2025, 112 (1), 182–188. DOI: 10.4269/ajtmh.23-0907.

(86) Kocalar, S.; Miller, B. M.; Huang, A.; Gleason, E.; Martin, K.; Foley, K.; Copeland, D. S.; Jewett, M. C.; Saavedra, E. A.; Kraves, S. Validation of Cell-Free Protein Synthesis Aboard the International Space Station. ACS Synth Biol 2024, 13 (3), 942–950. DOI: 10.1021/acssynbio.3c00733.

(87) FDA. Commercially Distributed Analyte Specific Reagents (ASRs). 2007.

(88) Boles, K. S.; Kannan, K.; Gill, J.; Felderman, M.; Gouvis, H.; Hubby, B.; Kamrud, K. I.; Venter, J. C.; Gibson, D. G. Digital-to-biological converter for on-demand production of biologics. Nat Biotechnol 2017, 35 (7), 672–675. DOI: 10.1038/nbt.3859.

(89) Kosuri, S.; Eroshenko, N.; Leproust, E. M.; Super, M.; Way, J.; Li, J. B.; Church, G. M. Scalable gene synthesis by selective amplification of DNA pools from high-fidelity microchips. Nat Biotechnol 2010, 28 (12), 1295–1299. DOI: 10.1038/nbt.1716.

(90) Espah Borujeni, A.; Channarasappa, A. S.; Salis, H. M. Translation rate is controlled by coupled trade-offs between site accessibility, selective RNA unfolding and sliding at upstream standby sites. Nucleic Acids Res 2014, 42 (4), 2646–2659. DOI: 10.1093/nar/gkt1139.

(91) Norouzi, M.; Panfilov, S.; Pardee, K. High-Efficiency Protection of Linear DNA in Cell-Free Extracts from. ACS Synth Biol 2021, 10 (7), 1615–1624. DOI: 10.1021/acssynbio.1c00110.

(92) Levine, M. Z.; Gregorio, N. E.; Jewett, M. C.; Watts, K. R.; Oza, J. P. Escherichia coli-Based Cell-Free Protein Synthesis: Protocols for a robust, flexible, and accessible platform technology. J Vis Exp 2019, (144). DOI: 10.3791/58882.

(93) Caschera, F.; Noireaux, V. Preparation of amino acid mixtures for cell-free expression systems. Biotechniques 2015, 58 (1), 40–43. DOI: 10.2144/000114249.

(94) Rupprecht, K. R.; Nair, R. K.; Harwick, L. C.; Grote, J.; Beligere, G. S.; Rege, S. D.; Chen, Y. Y.; Lin, Z.; Fishpaugh, J. R. Development of a dot-blot assay for screening monoclonal antibodies to low-molecular-mass drugs. Anal Biochem 2010, 407 (2), 160–164. DOI: 10.1016/j.ab.2010.08.003.

